# Genomics of chronic cough unravels neurological pathways

**DOI:** 10.1101/2024.07.23.24310853

**Authors:** Kayesha Coley, Catherine John, Jonas Ghouse, David J. Shepherd, Nick Shrine, Abril G. Izquierdo, Stavroula Kanoni, Emma F. Magavern, Richard Packer, Lorcan McGarvey, Jaclyn A. Smith, Henning Bundgaard, Sisse R. Ostrowski, Christian Erikstrup, Ole B. V. Pedersen, David A. van Heel, Genes & Health Research Team, William Hennah, Mikko Marttila, Robert C. Free, Edward J. Hollox, Louise V. Wain, Martin D. Tobin, Chiara Batini

**Affiliations:** Department of Population Health Sciences, University of Leicester, Leicester, UK; Laboratory for Molecular Cardiology, Department of Cardiology, Copenhagen University Hospital, Rigshospitalet, Building 9312, Henrik Harpestrengs Vej 4C, 2100 Copenhagen, Denmark; Laboratory for Molecular Cardiology, Department of Biomedical Sciences, University of Copenhagen, Copenhagen, Denmark; University Hospitals of Leicester NHS Trust, Groby Road, Leicester LE3 9QP, UK; William Harvey Research Institute, Barts and the London School of Medicine and Dentistry, Queen Mary University of London, London, UK; Wellcome-Wolfson Institute for Experimental Medicine, School of Medicine, Dentistry and Biomedical Sciences, Queen’s University Belfast, Belfast, UK; Division of Immunology, Immunity to Infection and Respiratory Medicine, The University of Manchester, Manchester University NHS Foundation Trust, Manchester, UK; Department of Cardiology, Copenhagen University Hospital, Rigshospitalet, University of Copenhagen, Copenhagen, Denmark; Department of Clinical Medicine, University of Copenhagen, Copenhagen, Denmark; Department of Clinical Immunology, Rigshospitalet, Copenhagen University Hospital, Copenhagen, Denmark; Department of Clinical Immunology, Aarhus University Hospital, Aarhus, Denmark; Department of Clinical Medicine, Aarhus University, Aarhus, Denmark; Department of Clinical Immunology, Zealand University Hospital, Køge, Denmark; Blizard Institute, Barts and the London School of Medicine and Dentistry, Queen Mary University of London, London, UK; Orion Pharma, Espoo, Finland; Neuroscience Center, HiLIFE, University of Helsinki, Helsinki, Finland; Orion Pharma, Nottingham, UK; School of Computing and Mathematical Sciences, University of Leicester, Leicester, UK; Department of Genetics and Genome Biology, University of Leicester, Leicester, UK

## Abstract

**Background:** Chronic cough is a symptom of common lung conditions, occurs as a side effect of ACE inhibitors (ACEis), or may be unexplained. Despite chronic cough representing a substantial health burden, its biological mechanisms remain unclear. We hypothesised shared genetic architecture between chronic dry cough and ACEi-induced cough and aimed to identify causal genes underlying both phenotypes.

**Methods:** We performed multi-ancestry genome-wide association studies (GWAS) of chronic dry cough and ACEi-induced cough, and a multi-trait GWAS of both phenotypes, utilising data from five cohort studies. Chronic dry cough was defined by questionnaire responses, and ACEi-induced cough by treatment switches or clinical diagnosis in electronic health records. We mapped putative causal genes and performed phenome-wide association studies (PheWAS) of associated variants and genetic risk scores (GRS) for these phenotypes to identify pleotropic effects.

**Findings:** We found seven novel genetic association signals reaching *p*-value <5×10^−8^ in the multi-trait or single-trait analyses of chronic dry cough and ACEi-induced cough. The novel variants mapped to 10 novel genes, and we mapped an additional three novel genes to known risk variants, many of which implicating neurological functions (*CTNNA1, KCNA10, MAPKAP1, OR4C12, OR4C13, SIL1*). The GRS-PheWAS highlighted associations with increased risk of several conditions reported as comorbidities of chronic cough, including fibromyalgia pain, and with spirometry measurements.

**Interpretation:** Our findings advance the understanding of neuronal dysfunction underlying cough hypersensitivity in chronic dry cough and ACEi-induced cough at the population-level, and the identification of comorbidities associated with genetic predisposition to cough could inform drug target discovery.

**Funding:** Medical Research Council, Wellcome Trust, National Institute for Health and Care Research, Orion Pharma.

**Research in context:** *Evidence before this study:* We searched the National Human Genome Research Institute-European Bioinformatic Institute Catalog of human genome-wide association studies (GWAS) from inception through to 21^st^ May 2024, using the search term “cough” to identify publications which tested association between genetic variants and cough. Additionally, we searched PubMed for English language articles published before 21^st^ May 2024 using the terms “cough” or “ACE inhibitor” combined with “genome-wide association” to find relevant publications. We manually filtered the results from both searches to ensure the cough-related publications related to a dry, unproductive cough. To date, there have been no GWAS of unexplained dry cough, while several for ACE inhibitor (ACEi)-induced cough have identified 11 associated genetic loci at genome-wide significance (*p*-value <5×10^−8^). These loci have implicated genes involved in neuronal excitability and the bradykinin pathway.

*Added value of this study:* We present the first multi-ancestry GWAS of chronic dry cough, defined using questionnaire responses, and the largest multi-ancestry GWAS of ACEi-induced cough, characterised by either a medication switch from an ACEi to an angiotensin-II receptor blocker or a clinical diagnosis. By leveraging the clinical and genetic overlap between these two traits, we conducted a multi-trait GWAS to identify novel associated loci. Across the multi-trait and single-trait analyses, we identified seven novel associated loci at genome-wide significance (*p*-value <5×10^−8^), mapped to 10 genes which have not previously been linked to cough phenotypes.

*Implications of all the available evidence:* We identified novel genes involved in neurological processes (*CTNNA1, KCNA10, OR4C12, OR4C13*) or implicated in neurological or neurodegenerative conditions (*SIL1* and *MAPKAP1*). The genes we implicate from all genetic associations with cough to date support the role of neurobiological processes underlying dry cough biology, and highlight potential therapeutic targets.

## Introduction

Chronic cough is common, impairs quality of life and results in substantial healthcare system burden^1^. Despite this, its molecular causes remain unclear and treatments are under-developed^2^. It may occur due to lung diseases, other identifiable conditions (known as ‘treatable traits,) such as ACE inhibitor (ACEi) use, eosinophilic airway inflammation, smoking, obesity, gastro-oesophageal reflux, rhinitis and obstructive sleep apnoea, or remain unexplained with no identifiable comorbidities^3-6^. The term cough hypersensitivity syndrome has been used to describe troublesome coughing triggered by low levels of thermal, mechanical or chemical exposure^7^. Neuronal dysregulation has been postulated to underlie cough hypersensitivity, which is thought to be a distinct treatable trait^8^. Recent studies have demonstrated the efficacy of therapies targeting neuronal mechanisms in patients with features consistent with cough hypersensitivity, diagnosed with refractory or unexplained chronic cough^2,7^.

Genetic studies guide understanding of mechanisms and drug targets^9^ but whilst the genetics of asthma, chronic obstructive pulmonary disease and chronic sputum production have been well-studied^10-12^, there has been a lack of genome-wide association studies (GWAS) of chronic cough. In this study, we explore two cough phenotypes: chronic dry cough and ACEi-induced cough, both of which have similar clinical manifestations, including a dry nature and involvement of cough reflex hypersensitivity. There have been no GWAS of chronic dry cough and few for ACEi-induced cough^13-16^, with findings from the latter implicating neuronal excitability and the bradykinin pathway^13,16^. We hypothesised that there would be common genetic determinants of chronic dry cough and ACEi-induced cough, in which case a multi-trait GWAS increases power to detect shared associated genetic variants^17^.

Therefore we: (i) performed the first GWAS of chronic dry cough; (ii) conducted the largest multi-ancestry study of ACEi-induced cough; (iii) tested genetic correlation between chronic dry cough and ACEi-induced cough; (iv) undertook the first multi-trait GWAS of chronic dry cough and ACEi-induced cough; (v) implemented a consensus-based framework to investigate putative causal genes for these traits and (vi) applied phenome-wide association studies (PheWAS) to associated variants across single-trait and multi-trait GWAS, as well as a multi-trait weighted genetic risk score (GRS) to inform understanding of the potential consequences of perturbing pathways involved in chronic cough. Using these approaches, we detected novel genetic associations and identified causal genes and pathways underlying these under-studied cough phenotypes.

## Methods

### Study populations and phenotype definitions

Our study included individuals from UK Biobank^18^, EXCEED^19^ Genes & Health^20^ and Copenhagen Hospital Biobank^13,21^, and summary statistics from a previously published study conducted in the eMERGE Network^16,22^ (further cohort details provided in **Appendix pp 2**).

We defined ACEi-induced cough using electronic health records (EHRs) linked to UK Biobank^18^, EXCEED^19^, Genes & Health^20^ and Copenhagen Hospital Biobank^13,21^ whereby cases switched from an ACEi to an angiotensin-II receptor blocker (ARB) and controls were continuous users of ACEis. The eMERGE Network study defined ACEi-induced cough using a validated algorithm based on prescriptions and the allergy section in EHRs^16,22^ (**Appendix pp 2**).

We defined chronic dry cough in UK Biobank^18^ using questionnaire data. Among UK Biobank participants who were neither ACEi-induced cough cases or controls, we defined chronic dry cough cases (non-productive cough on most days) and controls who neither coughed on most days nor brought up sputum on most days (**Appendix pp 2**).

### Genome-wide association studies

We performed ancestry-specific GWAS of chronic dry cough and ACEi-induced cough using imputed genomic variants (imputation quality _≥_0.3, and minor allele count [MAC] _≥_10), and age, age^2^, sex, genotyping array and at least 10 principal components of genetic ancestry as covariates. For chronic dry cough, ever-smoking status was an additional covariate. We also utilised summary statistics from the previously published eMERGE Network study^16^ (**Appendix pp 2,5-6**).

Using a fixed effects inverse variance-weighted model^23^ we undertook a GWAS meta-analysis for chronic dry cough and a GWAS meta-analysis for ACEi-induced cough (**Figure 1; Appendix pp 2**). Then we conducted a multi-trait GWAS^17^ by meta-analysis of the chronic dry cough GWAS and ACEi-induced cough GWAS using a fixed effects inverse-variance weighted model^23^ (**Figure 1; Appendix pp 2**). To inform the GRS development detailed below, we also performed a European (EUR)-only multi-trait GWAS.

**Figure 1.**
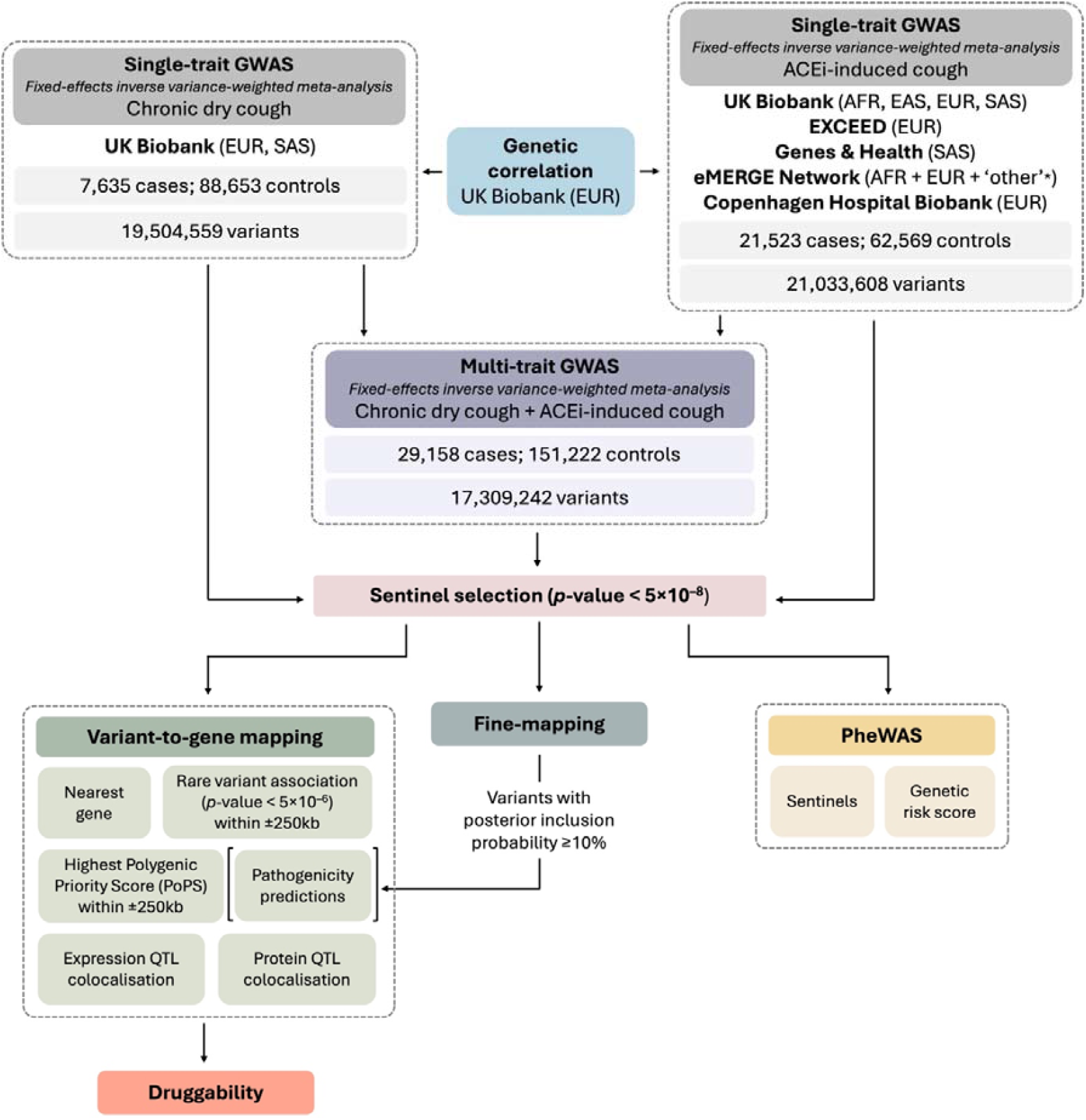
Study design. *Abbreviations:* ACEi, ACE inhibitor; AFR, African; EAS, East Asian; EUR, European; PheWAS, phenome-wide association study; SAS, South Asian; QTL, quantitative trait loci. *Defined by study authors^16^.

**Figure 2.**
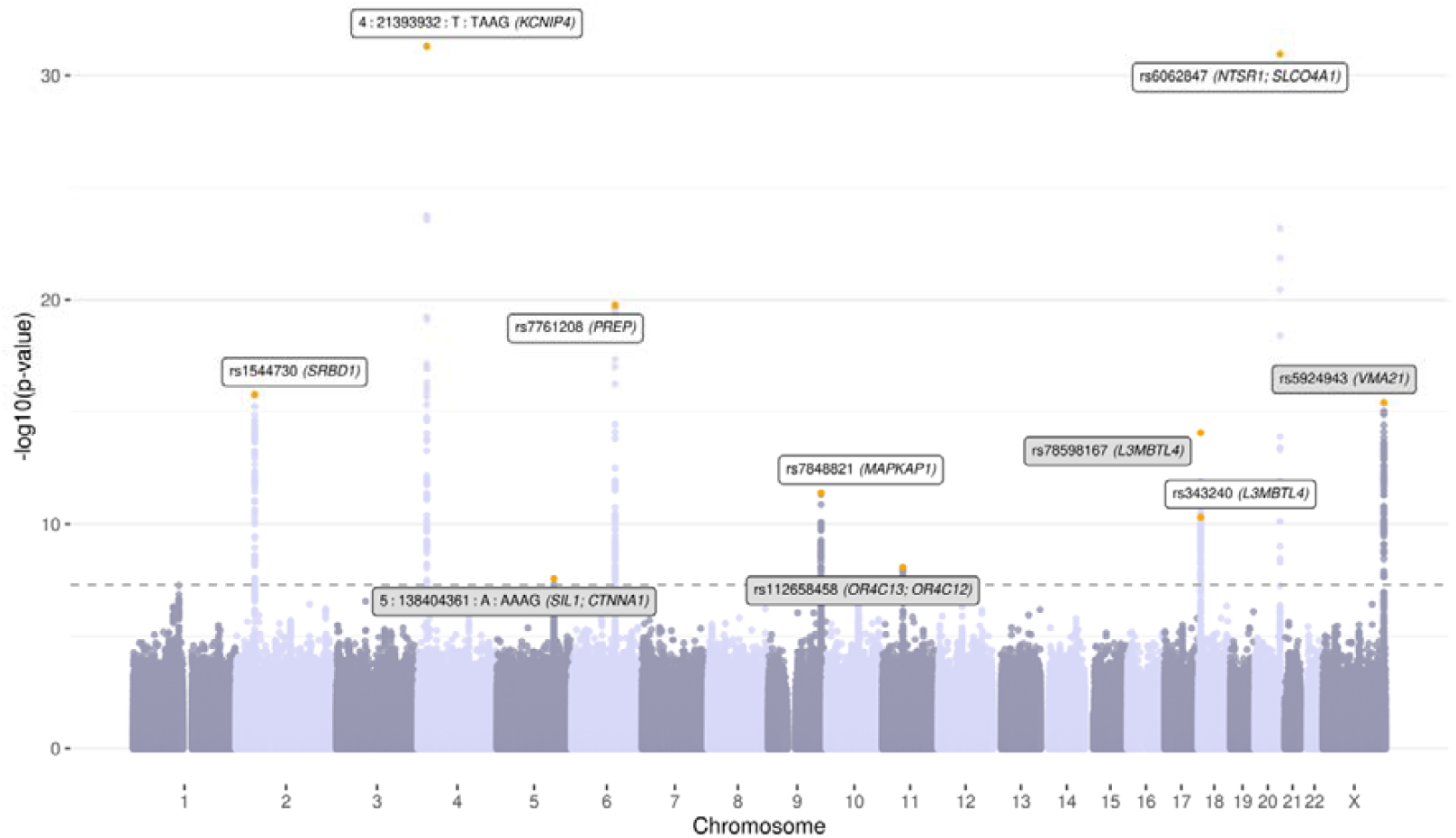
Manhattan plot for the multi-trait GWAS of chronic dry cough and ACEi-induced cough. The dotted grey line represents the genome-wide significance threshold (*p*-value <5×10^−8^). Sentinels are highlighted orange and mapped genes are labelled; novel sentinels are highlighted grey.

In each GWAS meta-analysis, we defined sentinel variants as those reaching genome-wide significance (*p*-value <5×10^−8^) with the lowest *p*-value within a ±1Mb locus. We identified secondary sentinels (*p*-value <5×10^−8^) within each locus using conditional analyses in each ancestry group using GCTA^24,25^ (**Appendix pp 3**). We fine-mapped each sentinel locus to calculate the posterior inclusion probability (PIP) for each variant, obtaining sets of variants with a 95% probability of containing a causal variant, referred to as ‘credible sets, (**Appendix pp 3**).

We defined single-trait sentinels as independent from multi-trait sentinels based on a linkage disequilibrium (LD) threshold of *r*^*2*^ <0.1. We also used *r*^*2*^ <0.1 to define novel multi-trait and single-trait (ACEi-induced cough) sentinels from previously reported associations (*p*-value <5×10^−8^) for ACEi-induced cough^13,15,16^, given the absence of published GWAS of chronic dry cough.

We estimated SNP heritability of chronic dry cough and of ACEi-induced cough and genetic correlation between these traits using LD Score Regression (LDSC)^26,27^ in European (EUR) individuals (**Appendix pp 3**).

### Investigating clinical and biological relevance of genetic associations

We mapped genes to all sentinel variants, both novel and previously reported, by integrating the following evidence: (i) nearest gene; (ii) a credible set containing a deleterious or damaging variant with a PIP _≥_10%; (iii) the gene with the highest Polygenic Priority Score (PoPS)^28^ within ±250kb of the sentinel for the relevant trait (multi-trait, chronic dry cough or ACEi-induced cough); (iv) exome-wide rare variant association (*p*-value <5×10^−6^) with the relevant trait in UK Biobank within ±250kb of the sentinel; (v) association with gene expression and (vi) protein expression (**Appendix pp 3-4**). Mapped genes supported by at least one level of evidence were considered putative causal, and we used database (GeneCards^29^, Open Targets Platform^30^, Online Mendelian Inheritance in Man^31^ [OMIM], Drug Gene Interaction Database [DGIdb]^32^) and literature searches to retrieve information about their biological function and clinical relevance.

We calculated GRS in UK Biobank EUR individuals, comprising all sentinel variants, each weighted by the estimated effect in the EUR-only multi-trait GWAS of chronic dry cough and ACEi-induced cough (outlined above; **Appendix pp 4**). We used DeepPheWAS^33^ to test association between sentinels individually (across multiple ancestries) and collectively via the GRS (in EUR only) and 1,939 phenotypes defined in UK Biobank, highlighting associations with a false discovery rate (FDR) <0.01. This was complemented by a custom Respiratory PheWAS to query variants against the most powerful GWASs performed across ten clinical respiratory traits (**Appendix pp 4,16**) and identifying associations with a *p*-value <0.001, and by querying Open Targets Genetics^34^ to assess whether sentinels were in LD (*r*^*2*^ >0.8) with genome-wide significant lead GWAS variants for other traits.

### Sensitivity analysis

We performed a sensitivity analysis in UK Biobank excluding individuals with asthma (defined by DeepPheWAS^33^ using both self-report and EHR data). For our sentinel variants, we followed the primary analysis protocol (outlined above; **Appendix pp 2,5-6**) and performed ancestry-specific association testing with chronic dry cough and ACEi-induced cough, excluding asthma cases. Using an inverse-variance weighted fixed effects model, we conducted separate meta-analyses for each trait, followed by a combined multi-trait meta-analysis of both traits.

## Results

The multi-trait GWAS of chronic dry cough and ACEi-induced cough included 29,158 cases and 151,222 controls, comprising 7,635 cases and 88,653 controls for chronic dry cough, and 21,523 cases and 62,569 controls for ACEi-induced cough (**Figure 1**; **Appendix pp 6**). SNP heritability for chronic dry cough was 3.19% (SE = 0.53%) and for ACEi-induced cough was 5.30% (SE 1.4%) and there was a strong genetic correlation between chronic dry cough and ACEi-induced cough (*r*_*g*_ = 0.56, SE = 0.15, *p*-value = 0.0001). Across the multi-trait and single-trait analyses, we defined 14 independent signals, identified by sentinel variants reaching *p*-value <5×10^−8^ (**Appendix pp 7-8**). These signals mapped to a total of 19 genes through our comprehensive variant-to-gene mapping approach (**Figure 3; Appendix pp 9-12,25**), and were subsequently explored for gene-drug interactions (**Appendix pp 13**).

**Figure 3.**
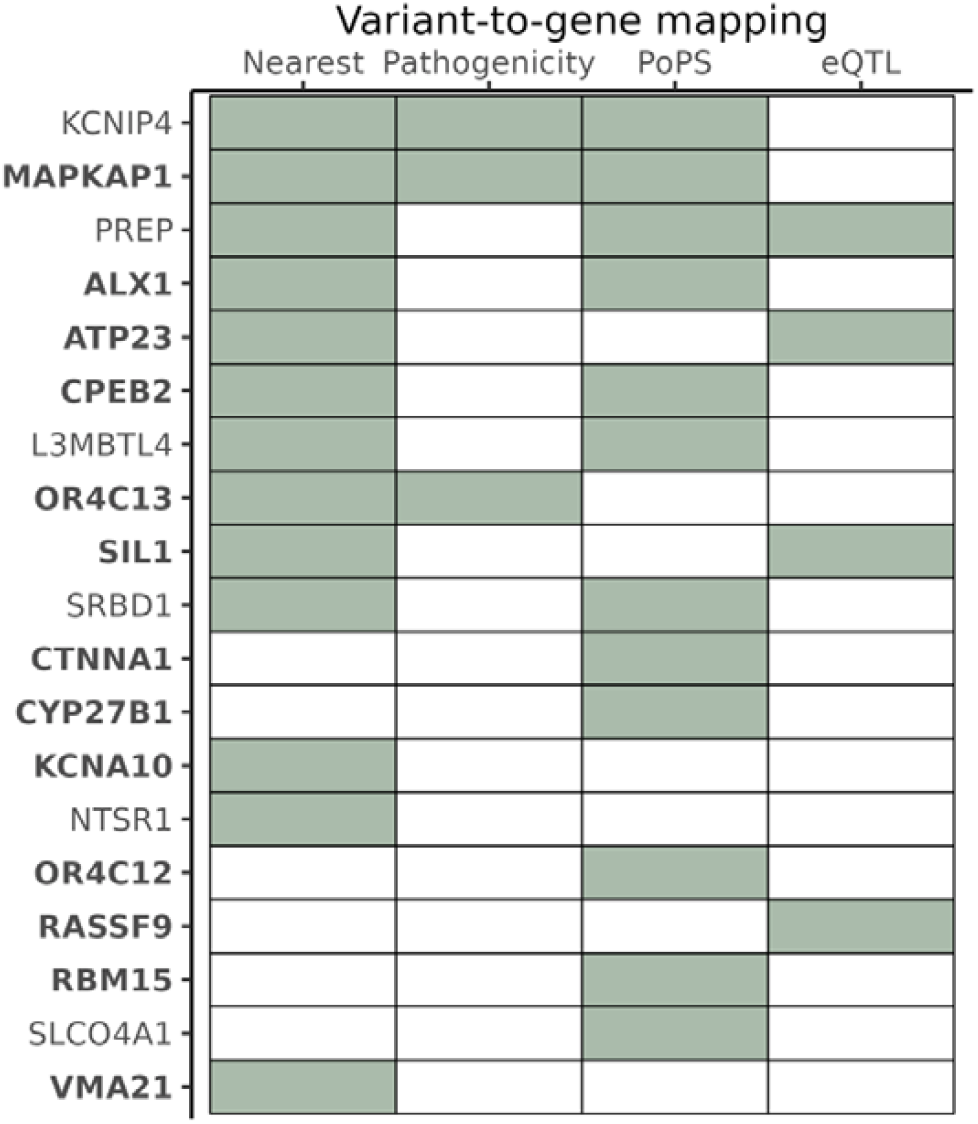
Genes identified by variant-to-gene mapping. Filled tiles indicate genes identified by specific analysis. Protein quantitative trait loci (pQTL) and rare variant columns have been omitted as these did not lead to the mapping of any genes. Novel genes are highlighted bold. *Abbreviations:* eQTL, expression quantitative trait loci; PoPS, Polygenic Priority Score; pQTL, protein quantitative trait loci.

We found seven novel genetic association signals in the multi-trait or single-trait analyses of chronic dry cough and ACEi-induced cough (**Table 1**, see **Appendix pp 7-8** for further details). In the multi-trait GWAS, four novel signals mapped to six genes, five of which (*CTNNA1, OR4C12, OR4C13, SIL1* and *VMA21*) have not been previously implicated in any cough GWAS (**Figure 2**; **Figure 3**). The remaining gene, *L3MBTL4*, was previously implicated by a common variant (rs8097200) associated with ACEi-induced cough^13^, and independent from our sentinel, rs78598167 (*r*^2^ = 0.003).

**Table 1.**
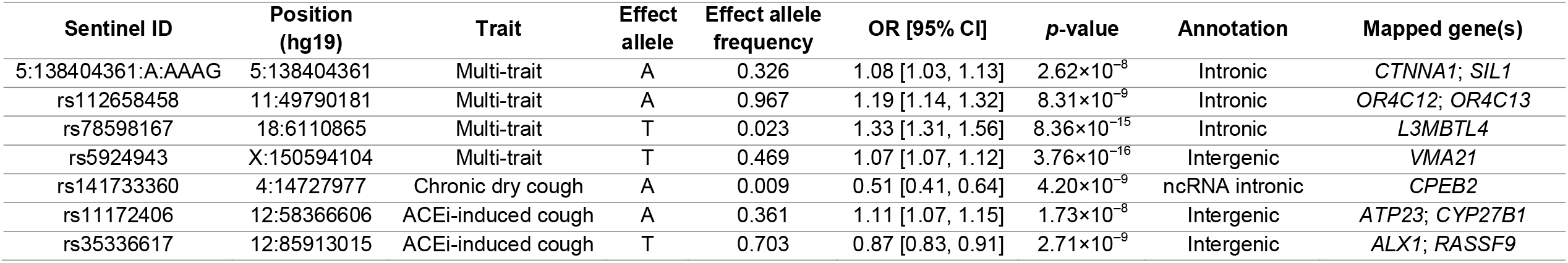
Novel sentinel variants (*p*-value <5×10^−8^) identified in the multi-trait GWAS of chronic dry cough and ACEi-induced cough, and additional novel sentinels from each single-trait GWAS. *Abbreviations:* CI, confidence interval; OR, odds ratio.

The single-trait analyses identified three novel signals mapping to 5 novel genes (**Table 1**). While the sentinel variant found for chronic dry cough (rs141733360), and its mapped gene (*CPEB2*), have not been previously described in the literature as associated with any cough phenotype, the only phenotypes we observed associated with rs141733360 in the PheWAS were cough-related (**Appendix pp 14**). The two sentinel variants for ACEi-induced cough mapped to *ATP23* and *CYP27B1* (rs11172406), and *ALX1* and *RASSF9* (rs35336617), none of which have been identified before for any cough phenotype. Our PheWAS showed that the allele associated with increased ACEi-induced cough (allele A of rs11172406), mapping to *ATP23* and *CYP27B1*, was associated with reduced urea and urate levels. This finding was supported by the Open Targets query for its proxy rs10877067 (in high LD with rs11172406, *r*^2^ = 0.88); allele T at this variant which increased risk of ACEi-induced cough was also associated with reduced urate levels (**Appendix pp 14-15**).

We also identified seven previously reported signals for ACEi-induced cough, six in the multi-trait analysis and one in the ACEi-induced cough single-trait analysis. Collectively, these mapped to nine genes (*KCNA10, KCNIP4, L3MBTL4, MAPKAP1, NTSR1, PREP, RBM15, SLCO4A1* and *SRBD1*), three of which (*KCNA10, MAPKAP1* and *RBM15*) were never implicated before for any cough phenotype (**Appendix pp 7**). The PheWAS showed that the signal mapping to *MAPKAP1* was associated with increased risk of asthma^12^ (*p*-value = 3.70×10^−4^) [**Appendix pp 16-17**], while the signal mapping to *NTSR1* and *SLCO4A1* was associated with both risk of asthma^12^ (*p*-value = 1.70×10^−5^) [**Appendix pp 16-17**] and cough-related phenotypes (FDR <1%, **Appendix pp 14**) with the same direction of effect. However, the effect size estimates of all sentinel variants were not systematically or substantially attenuated after exclusion of asthma cases (**Appendix pp 18**).

We identified the gene product of *KCNA10* as a target for several potassium channel blockers, including fampridine, which has been approved for multiple sclerosis treatment, as well as others currently undergoing clinical trials for other indications including stroke and spinal cord injury (**Appendix pp 13**). Reminertant, a drug targeting the protein encoded by *NTSR1* is also undergoing trials for small cell lung carcinoma (**Appendix pp 13**).

Five of the seven sentinels previously reported for ACEi-induced cough^13^ showed association (*p*-value < 0.05) with chronic dry cough, more than expected by chance (binomial test *p*-value = 6×10^−6^). These mapped to the *SRBD1, KCNIP4, PREP, SCAI* and *NTSR1/SLCO4A1* genes^13^, all five variants showing a consistent direction of effect between chronic dry cough and ACEi-induced cough and a larger effect size estimate for chronic dry cough than for cough on most days^13^. The association with chronic dry cough met a Bonferroni threshold for the sentinels at the *SRBD1* (rs1544730), *PREP* (rs7761208), and *NTSR1/ SLCO4A1* (rs6062847) loci.

We developed a GRS for chronic cough based on the 14 sentinel variants weighted by the multi-trait analysis to maximise power for the PheWAS. Genetically predicted chronic cough was associated with quantitative phenotypes including lower forced expiratory volume in one second and lower forced vital capacity, lower fat free mass, reduced hand grip strength and lower urate levels (**Appendix pp 19-24**). Furthermore, an increase in the GRS was associated with disease risk, such as an increased risk of breast cancer and osteoporosis, and reduced risk of gout, diabetes and ischaemic heart disease. Most notably, an increase in the GRS was associated with increased risk of migraine, irritable bowel syndrome (IBS), fibromyalgia pain, hypothyroidism and urinary incontinence – all of which have been reported as comorbidities accompanying chronic cough^35-38^ (**Appendix pp 19-24,26**).

## Discussion

Chronic cough is a common symptom with prevalence estimates varying from 1-2% up to 10%^4,39^, and imposes a substantial health burden^1^. Despite this, its biological mechanisms are not fully understood. We have utilised GWAS approaches to identify associated genetic variants and implicate relevant genes, shedding light on the molecular basis of chronic dry cough and ACEi-induced cough, and providing genetic evidence for future therapeutic developments. Specifically, we undertook the first GWAS of chronic dry cough and the largest multi-ancestry GWAS of ACEi-induced cough to date, and having demonstrated genetic overlap between chronic dry cough and ACEi-induced cough, we performed a multi-trait GWAS, thereby maximising power to discover novel variants associated with both traits. Further, we showed that collectively, cough-associated variants were associated with raised risk of conditions such as migraine, IBS, fibromyalgia pain, hypothyroidism, urinary incontinence, breast cancer, and osteoporosis, providing further insights as to the potential consequences of modulation of pathways involved in chronic cough.

We found seven novel sentinel variants associated with these cough phenotypes which reached *p*-value <5×10^−8^ in the multi-trait or single-trait analyses and highlight the relevance for chronic dry cough of five further sentinels previously reported for ACEi-induced cough. The seven novel sentinels mapped to 10 genes not previously implicated in cough phenotypes (*SIL1, CTNNA1, OR4C12, OR4C13, VMA21, CPEB2, ATP23, CYP27B1, ALX1, RASSF9*), and through a comprehensive variant-to-gene mapping strategy we additionally mapped known loci to a further three novel genes (*MAPKAP1, KCNA10, RMB15*). Descriptions of novel (and previously reported) genes are provided in **Appendix pp 25**.

Collectively, the novel sentinels and genes, alongside those previously reported, enrich our understanding of the biological mechanisms underlying chronic dry cough and ACEi-induced cough. Many of the novel putative causal genes encode proteins with neurological functions. CTNNA1 is a member of the catenin protein family which is involved in cell-cell adhesion^29^, synapse morphogenesis and plasticity^40^. *KCNA10* encodes a subunit of potassium ion channels (as does *KCNIP4*^*13*^ highlighted below) which have a role in neuronal excitability, neurotransmitter release and smooth muscle contraction^29,30^, and is a target for an approved drug for symptom management in multiple sclerosis. MAPKAP1 is a subunit of the mTOR complex 2, a component of the mTOR signalling pathway vital for growth and metabolism^29,30,41^. *MAPKAP1* has been identified in GWAS of pain intensity^42^, and dysregulation of the mTOR signalling pathway has been implicated in various neurodegenerative and neuropsychiatric disorders, including epilepsy^43^. OR4C12 and OR4C13 are G-protein coupled receptors predominantly expressed in nasal epithelium olfactory sensory neurons and involved in neural response to odorant stimuli^29,30^. SIL1 is a glycoprotein involved in protein translocation and folding and is associated with Marinesco-Sjogren syndrome^31^ which is characterised by a number of neurological symptoms such as cerebellar ataxia and intellectual disability^44^.

Further, we provide evidence that loci, genes and pathways previously implicated in ACEi-induced cough have a role in chronic dry cough, and may provide insights into potential therapeutic targets^13,16,45^. The genes previously implicated for ACEi-induced cough also highlight the importance of neurological processes. In particular, PREP is involved in the mediation of neuropeptide activity, including the metabolism of bradykinin^46,47^ which has been shown to sensitise airway sensory nerves and induce cough reflex hypersensitivity in an animal model via the activation of airway C-fibres^48^. *KCNIP4* modulates neuronal excitability^29^, while NTSR1 mediates the activity of neurotensin, a pro-inflammatory peptide involved in the modulation of smooth muscle contraction^49^, and also upregulates the secretion of GABA^30^. Animal models and controlled trials have demonstrated that centrally acting agonists of GABA_B_ receptors inhibit the cough reflex; with baclofen being reported to suppress ACEi-induced cough^50,51^. Additionally, animal studies have provided evidence for the role of neuronal potassium channels in smooth muscle reactivity and cough reflex modulation^52^.

We found that GRSs for our cough phenotypes were associated with increased risk of several recognized comorbidities of chronic cough, including fibromyalgia pain. The neurobiological mechanisms of chronic cough and chronic pain exhibit similarities, with central and peripheral sensitization contributing to their underlying symptoms, and afferent nerve fibers involved in both conditions expressing common receptors^53^. P2X3 receptors are involved in nerve fiber sensitization in chronic pain^54^, and P2X3 receptor antagonists have shown antitussive efficacy in refractory and unexplained chronic cough patients in clinical trials^55^.

By leveraging questionnaire responses from UK Biobank to define chronic dry cough and primary care EHR prescribing data to characterise ACEi-induced cough, we conducted the most powerful GWAS of chronic cough to date. This phenotyping approach allowed us to maximise the use of relevant phenotypic data available and overcome the limitation that drug response and adverse reaction-related traits are rarely specifically recorded in study populations, facilitating the study of ACEi-induced cough in large sample sizes across multiple cohorts. Another strength of this study is the implementation of a detailed variant-to-gene mapping framework to implicate putative causal genes and biological mechanisms. Further, we utilised recently developed techniques to improve power for understanding the potential consequences of pathway modulation through GRS-informed PheWAS. We also note limitations of our study. Although we included individuals of diverse ancestries, the sample sizes for non-European groups remained modest despite our efforts to maximise data availability by utilising both EHRs and questionnaire responses. This bias presents an important issue for many GWASs^56^ and highlights the importance of initiatives with a recruitment focus on underrepresented populations to improve generalisability of findings across populations^57^. Additionally, as with any case-control study, misclassification of cases and controls is possible, perhaps more so using an ACEi to ARB switch to characterise cases. However, such misclassification would tend to reduce effect size estimates towards the null and potentially miss some true associations, and an ACEi to ARB switch to capture ACEi-induced cough has been validated through manual interrogation of patient histories^15^, consistency of genetic associations^13,16^ and polygenic risk score predictions^13^. We note that variable terms are used to describe and categorise chronic cough, including cough hypersensitivity syndrome, refractory chronic cough and unexplained chronic cough^58^. We did not attempt to distinguish these conditions, which would generally require specialist clinical assessment given that cough is poorly coded in EHRs^6^ and not widely characterised in large cohort questionnaires. Such studies will require further development of registries of well-characterised patients with samples and consent for genetic studies, such as NEuroCOUGH^2^.

Our findings expand on the established concept of cough hypersensitivity due to neuronal dysfunction, which in specific cases can be identified as a treatable trait, by showing that this mechanism applies broadly to chronic dry cough at the population-level, and to ACEi-induced cough. Further, identifying the association between genetic predisposition to cough with various known comorbidities, including chronic pain, could further advance our understanding of involved biological processes and should be considered in drug discovery and development efforts.

## Data Availability

Access to UK Biobank (https://www.ukbiobank.ac.uk), EXCEED (https://exceed.org.uk/), Genes & Health (https://www.genesandhealth.org/) and Copenhagen Hospital Biobank individual-level data are available to approved researchers upon application or data access request. Genome-wide summary statistics from the single-trait and multi-trait analyses will be made publicly available via the EMBL-EBI GWAS Catalog. The ACEi-induced cough phenotyping algorithm is publicly available (https://doi.org/10.5281/zenodo.6780065), and scripts used to run additional analyses are available upon request.

## Acknowledgements

We thank all volunteers participating in the EXCEED Study, UK Biobank, Genes & Health, Copenhagen Hospital Biobank and the eMERGE Network and who have made this project possible. We also thank authors of the eMERGE Network study for sharing GWAS summary statistics. This study used the ALICE High Performance Computing Facilities at the University of Leicester.

## Copenhagen Hospital Biobank

The analysis was performed under the Copenhagen Hospital Biobank Cardiovascular Disease Cohort (CHB-CVDC), approval number NVK-1708829, P-2019-93.

### EXCEED

EXCEED has been supported by the University of Leicester, the National Institute for Health and Care Research Leicester Respiratory Biomedical Research Centre, the Wellcome Trust (WT 202849), and Medical Research Council grants G0902313 and UKRI_PC_19004, the latter through the UK Research and Innovation Industrial Strategy Challenge Fund, delivered through Health Data Research UK. EXCEED has also been supported by Cohort Access fees from studies funded by the Medical Research Council (MRC), Biotechnology and Biological Sciences Research Council, National Institute for Health and Care Research, the UK Space Agency, and GlaxoSmithKline. EXCEED received ethical approval from the Leicester Central Research Ethics Committee (13/EM/0226).

Substantial amendments have been approved by the same Research Ethics Committee for the collection of new data relating to the COVID-19 pandemic, including the COVID-19 questionnaires and antibody testing.

### Genes and Health

Genes & Health is/has recently been core-funded by Wellcome (WT102627, WT210561), the Medical Research Council (UK) (M009017, MR/X009777/1, MR/X009920/1), Higher Education Funding Council for England Catalyst, Barts Charity (845/1796), Health Data Research UK (for London substantive site), and research delivery support from the NHS National Institute for Health Research Clinical Research Network (North Thames). Genes & Health is/has recently been funded by Alnylam Pharmaceuticals, Genomics PLC; and a Life Sciences Industry Consortium of Astra Zeneca PLC, Bristol-Myers Squibb Company, GlaxoSmithKline Research and Development Limited, Maze Therapeutics Inc, Merck Sharp & Dohme LLC, Novo Nordisk A/S, Pfizer Inc, Takeda Development Centre Americas Inc. We thank Social Action for Health, Centre of The Cell, members of our Community Advisory Group, and staff who have recruited and collected data from volunteers. We thank the NIHR National Biosample Centre (UK Biocentre), the Social Genetic & Developmental Psychiatry Centre (King,s College London), Wellcome Sanger Institute, and Broad Institute for sample processing, genotyping, sequencing and variant annotation. We thank: Barts Health NHS Trust, NHS Clinical Commissioning Groups (City and Hackney, Waltham Forest, Tower Hamlets, Newham, Redbridge, Havering, Barking and Dagenham), East London NHS Foundation Trust, Bradford Teaching Hospitals NHS Foundation Trust, Public Health England (especially David Wyllie), Discovery Data Service/Endeavour Health Charitable Trust (especially David Stables), Voror Health Technologies Ltd (especially Sophie Don), NHS England (for what was NHS Digital) - for GDPR-compliant data sharing backed by individual written informed consent.

### UK Biobank

UK Biobank is generously supported by its founding funders the Wellcome Trust and UK Medical Research Council, as well as the Department of Health, Scottish Government, the Northwest Regional Development Agency, British Heart Foundation and Cancer Research UK. UK Biobank genetic and phenotypic data were obtained under UK Biobank applications 43027 and 59822. Analyses of sequencing data were conducted on the Research Analysis Platform (https://ukbiobank.dnanexus.com). UK Biobank has ethical approval from the UK National Health Service (NHS) National Research Ethics Service (11/NW/0382).

For the purpose of open access, the author has applied a CC BY public copyright licence to any Author Accepted Manuscript version arising from this submission.

## Author contributions

Conceptualization: MDT and CB; Methodology: KC, DJS, MDT and CB; Software: KC, RP; Validation: KC; Formal Analysis and Interpretation: KC, JG, NS, AGI, SK, EFM, RP, WH, MM; Investigation: KC, CJ, MDT and CB; Resources: CJ, DJS, RP, HB, SRO, CE, OBVP, DAH, Genes & Health Research Team, RCF, EJH, LVW and MDT; Data Curation: KC, CB; Writing – Original Draft: KC, CJ, MDT, and CB; Writing – Review & Editing: all authors; Visualization: KC, NS; Supervision: MDT and CB; Funding Acquisition: EJH, LVW, MDT and CB.

