## Supplementary Material (Appendix) for "Genomics of chronic cough unravels neurological pathways"

**Supplementary Appendix**

[Supplementary Table 10. Sentinel variant PheWAS results (false discovery rate [FDR] <0.01) using DeepPheWAS.	14](#_Toc172120204)

[Supplementary Table 13. Sentinel variant Respiratory PheWAS results (*p*-value <0.001).	17](#_Toc172120207)

[Supplementary Table 15. Genetic risk score PheWAS results (false discovery rate [FDR] <0.01) using DeepPheWAS.	19](#_Toc172120209)

[Supplementary Figure 1. Genetic risk score-based PheWAS results (false discovery rate [FDR <0.01]) using DeepPheWAS, restricted to binary phenotypes.	26](#_Toc172120212)

### Supplementary Text

#### Study populations

UK Biobank^1^ is a large-scale resource of approximately 500,000 individuals from across the UK, aged 40-69 during the recruitment period between 2006 and 2010. The genotyping and imputation protocol for UK Biobank samples is described extensively in Bycroft *et al* (2018). Genetically-determined ancestry was defined using *k*-means clustering of the first two genetic principal components (PCs)^2^, which were provided by UK Biobank^1^.

The EXCEED Study^3^ is a cohort study with participants recruited from Leicester City, Leicestershire and Rutland. Driven mainly through general practices, recruitment started in 2013 and was aimed at individuals between 40 and 69 years. EXCEED samples were genotyped using the UK Biobank Axiom array, and samples were excluded prior to imputation where the call rate was <97% of if genetic sex did not match phenotypic sex. Variants with call rate <95%, Hardy-Weinberg *p*-value <1×10^–6^ or minor allele frequency (MAF) <1% were excluded. Genotype data were phased using Eagle v2.4 and imputation was performed using the TOPMed imputation server (imputation: Minimac4, reference panel: TOPMed r2). Genetic ancestry was defined using *k*-means clustering of the first two PCs (calculated using EIGENSOFT^4^) with 1000 Genomes^5^ as a reference.

Genes & Health^6^ (is a cohort of over 60,000 British Bangladeshi and British Pakistani individuals who have been recruited from across Bradford, East London, and Manchester since 2015. Further details are available in Finer *et al* (2020). Genotyping of Genes & Health samples was performed using Illumina GSAv3EAMD and imputed to the TOPMed reference panel.

Copenhagen Hospital Biobank^7^ is a Danish biobank established to leverage residual blood samples collected during patient hospitalisations. Genotyping of samples in Copenhagen Hospital Biobank was performed using the Illumina Global Screening Array and underwent standard quality control protocol described in Sorensen *et al* (2021). Genotyped data were phased using Eagle v2 and imputed to reference panel backbone consisting of whole-genome sequence data from 8,429 Danes and 7,146 samples from North-Western Europe^8^.

The Mosley *et al* study^9^ utilised the electronic Medical Records and Genomics (eMERGE) Network^10^, a collection of research institutions and medical centres across the United States which focusses on integrating biorepositories and electronic health records (EHRs). Genotyping was performed using various Illumina arrays (**Supplementary Table 1**), and quality control performed as described in Mosley *et al* (2016). Before imputing the genotype data to the 1000 Genomes phase 3^5^ reference panel using IMPUTE2, phasing was performed using SHAPEIT^9^. Genetic ancestry was defined using STRUCTURE with HapMap as a reference panel^9^.

#### Phenotype definitions

ACEi-induced cough was defined using primary care EHRs linked to UK Biobank^1^, EXCEED^3^ and Genes & Health^6^ participants. Cases switched from an ACEi to an ARB within 12 months of the date of their first ACEi prescription (*index date*) and did not receive any ACEi prescriptions in 12 months after the switch date, and controls received at least one ACEi prescription and no ARB prescriptions within 12 months after *index date*. The definition in Copenhagen Hospital Biobank utilised dispensary data and was based on a switch to an ARB after continuous ACEi treatment^7,8^. In the published eMERGE Network study, ACEi-induced cough was defined using a validated algorithm based on prescriptions and the allergy section in EHRs^9^.

Chronic dry cough was defined in UK Biobank using questionnaire fields 22502 (‘cough on most days’) and 22504 (‘bring up phlegm/sputum/mucus on most days’), as in the phenotype generation protocol implemented in DeepPheWAS^11^. Cases had a non-productive cough on most days (22502 = 1 & 22504 = 0) and controls did not have a productive cough on most days (22502 = 0 & 22504 = 0). We excluded individuals already defined as either ACEi-induced cough cases or controls, and any individuals related at second degree or closer to any ACEi-induced cough cases or controls.

#### Genome-wide association studies

The genome-wide association studies (GWASs) of chronic dry cough and ACEi-induced cough were performed under an additive genetic model using imputed genomic variants (imputation quality ≥ 0.3, and minor allele count [MAC] ≥10) in genetically-defined ancestral groups containing ≥30 cases (**Supplementary Table 1**). We also utilised summary statistics with MAF >1% and imputation quality ≥0.7 from the previously published study conducted in the eMERGE Network^9^ (**Supplementary Table 1**). All summary statistics were harmonised to genome build hg19 using *liftover*^12^. All ancestry-specific GWASs, single-trait GWASs, and the multi-trait GWAS had a genomic inflation factor (λ_GC_) <1.05, hence no genomic-control correction was applied.

Using a fixed effects inverse variance-weighted meta-analysis in METAL^13^, we meta-analysed chronic dry cough GWASs across UK Biobank European (EUR) and South Asian (SAS) groups. Similarly, we meta-analysed ACEi-induced cough GWASs undertaken in: UK Biobank (African [AFR], East Asian [EAS], EUR and SAS); EXCEED (EUR), Genes & Health (SAS), eMERGE Network (AFR, EUR, ‘other’, as defined by the Mosley *et al*)^9^ and Copenhagen Hospital Biobank (EUR). These single-trait meta-analyses were included in the multi-trait GWAS of chronic dry cough and ACEi-induced cough, this was performed using a fixed effects inverse variance-weighted model (in METAL^13^) and included genetic variants represented in both datasets.

We estimated SNP heritability of chronic dry cough and of ACEi-induced cough and genetic correlation between these traits using LD Score Regression (LDSC)^14,15^ with UK Biobank-EUR GWAS summary statistics from common HapMap3 variants and 1000 Genomes EUR derived LD Scores.

Conditionally independent signals were identified in each ancestral group separately using forward selection, followed by backward elimination in GCTA^16,17^. For each locus, unconditional summary statistics, and an ancestry matched reference panel (10,000 randomly selected EUR individuals in UK Biobank for EUR, or appropriate super-population from 1000 Genomes Phase 3^5^ for AFR, EAS, and SAS) were used as input. Firstly, in each ancestral group, we conditioned on each sentinel using --cojo-cond, meta-analysed across all ancestries (to compile each single-trait, and again to create the multi-trait). If the sentinel was not present in an ancestral group, this group was removed from the meta-analysis. If there were any variants with *p*-value <5×10^–8^, we ran another round of conditional analysis including the original sentinel, and this sentinel, and repeated the process until there were no more sentinels reaching *p*-value <5×10^–8^. In backward selection, we obtained joint *p*-values for all sentinels in the conditional set for each locus using --cojo-joint, meta-analysed across ancestries (to compile each single-trait, and again to create the multi-trait), and if any variant did not reach *p*-value <5×10^–8^ we removed the variant with the largest *p*-value from the conditional set. If required, we iterated between forward selection and backward elimination until no additional variants outside the conditional set were genome-wide significant in the meta-analysis, and all variants in the conditional set had *p*-value <5×10^–8^ in the joint model post meta-analysis.

We calculated LD between sentinel variants between the multi-trait sentinels and sentinels identified for each single trait, as well as all independent sentinels across the multi-trait and single-trait GWASs and previously reported variants, using the reference panel based on 10,000 randomly selected EUR individuals in UK Biobank. We used a threshold of *r*^2^ <0.1 to define independence and novelty.

Using the approximate Bayes Factor method^18^, we fine-mapped the 2Mb locus surrounding each sentinel variant calculate the posterior probability of inclusion (PIP) for each variant and generate 95% credible sets. We set the prior *W* to 0.04, and utilised unconditional summary statistics, unless a secondary signal was identified, then conditional summary statistics were used. We included variants with MAF >0.1% and *p*-value <0.001. After ranking variant-level PIPs from smallest to largest, the PIPs were summed until at least 95% was reached to create the 95% credible sets.

#### Investigating clinical and biological relevance of genetic associations

**(i) Nearest gene:** We identified nearby genes to each sentinel using ANNOVAR^19^ and selected the closest protein-coding gene.

**(ii) Variant annotation and pathogenicity estimates:** We used ANNOVAR and Variant Effect Predictor^20^ to annotate each variant with PIP ≥10%, obtained through fine-mapping of associated loci (described above) with its consequence, SIFT and PolyPhen-2 predictors, and Combined Annotation Dependent Depletion (CADD) score. The nearest genes to variants annotated as deleterious by SIFT, probably damaging by PolyPhen-2 or with a deleterious CADD Phred score of ≥12.37 were selected (**Supplementary Table 6**).

**(iii) Polygenic Priority Score (PoPS):** PoPS^21^ is a similarity-based method which leverages gene-level features derived from multiple sources, including gene expression datasets, biological pathways and protein-protein interactions, to perform gene prioritisation. We applied PoPS to the EUR-only summary statistics of the multi-trait GWAS and each single-trait GWAS to generate gene-level polygenic priority scores and used the EUR population from 1000 Genomes Project^5^ Phase 3 as an LD reference. For this analysis, we selected the gene with the highest polygenic priority score (based on results using the relevant summary statistics) within ±250kb of each variant (extended to ±500kb when no gene was identified within ±250kb) [**Supplementary Table 7**].

**(iv) Rare variant association:** To support identifying nearby rare (MAC ≥3 and MAF <1%) variant associations, we undertook exome-wide association studies (ExWASs) of chronic dry cough and ACEi-induced cough using the UK Biobank exome sequences (OQFE pipeline) and REGENIE^22^. The REGENIE protocol was the same as reported in Supplementary Table. GWAS protocols for participating cohorts, and we excluded variants which did not pass the following PLINK filters^23^: --geno 0.1 --hwe 1e-15 --mind 0.1. Using METAL^13^, for each trait we meta-analysed the ExWAS summary statistics from different ancestral groups, and subsequently meta-analysed both traits in a multi-trait ExWAS. Using the relevant ExWAS meta-analysis summary statistics, we selected genes harbouring a rare variant association (*p*-value <5×10^−6^) within ±250kb of each variant.

**(v) Expression quantitative trait loci (eQTL) colocalisation:** We performed a preliminary look-up of the lead variants in GWAS summary statistics of gene expression levels in two *cis*-eQTL resources: (i) eQTLGen^24^, a blood-only resource, and (ii) GTeX v8^25^, restricted to adrenal, artery, blood, brain, oesophagus, heart, ileum, liver, lung, muscle, nerve, pituitary, spleen and stomach tissues. We identified variant-gene-tissue combinations with *p*-value < 5×10^−8^, and tested the corresponding locus (in EUR only) for colocalisation with the eQTL signal using coloc.susie^26^. We used an LD reference based on imputed genotype data from 10,000 EUR individuals in UK Biobank. Genes with a H_4_ ≥0.7 were selected (**Supplementary Table 8**).

**(vi) Protein quantitative trait loci (pQTL) colocalisation:** We performed a preliminary look-up of the lead variants in GWAS summary statistics of protein levels across three pQTL resources: (i) deCODE Genetics^27^, (ii) the SCALLOP Consortium^28^; and (iii) UK Biobank-Pharma Proteomics Project (PPP)^29^. We identified variant-protein combinations with *p*-value < 1.8×10^−9^ in the deCODE Genetics dataset (as used to determine significance by the authors in the original publication^27^) or *p*-value <5×10^−8^ in the SCALLOP Consortium and UK Biobank-PPP datasets. For associated variants identified as *cis*-pQTLs, i.e. located within ±1Mb of the transcription start site of the gene encoding the measured protein, we tested the corresponding locus (in EUR only) for colocalisation with the pQTL signal using coloc.susie^26^. We used an LD reference based on imputed genotype data from 10,000 EUR individuals in UK Biobank. Encoding genes with a H_4_ ≥0.7 were selected.

We used the --score function in PLINK^30^ to construct a genetic risk score (GRS) from UK Biobank imputed genotypes using the effect sizes of the multi-trait sentinels in EUR-only multi-trait GWAS of chronic dry cough and ACEi-induced cough as weights.

For the Respiratory PheWAS, we performed a look-up of the sentinel variants in 26 GWASs performed across ten clinical respiratory traits: asthma, bronchiectasis, bronchopneumonia, chronic bronchitis, chronic obstructive pulmonary disease, chronic sputum production, emphysema, idiopathic pulmonary fibrosis, respiratory infections, and interstitial lung abnormality. The sources of these datasets are provided in **Supplementary Table 13**.

### Supplementary Tables

Supplementary Table 1. GWAS protocols for participating cohorts.
Abbreviations: AFR, African; EAS, East Asian; EUR, European; HRC, Haplotype Reference Consortium; MAC, minor allele count; MAF, minor allele frequency; PC, principal component; SAS, South Asian; TOPMed, Trans-Omics for Precision Medicine.

|  | **UK Biobank** | **EXCEED** | **Genes & Health** | **eMERGE Network  (Mosley *et al,* 2016)** | **Copenhagen Hospital Biobank** |
| --- | --- | --- | --- | --- | --- |
| **Genetic data** | | | | | |
| **Genotyping array** | Applied Biosystems UK BiLEVE Axiom; Applied Biosystems UK Biobank Axiom | Applied Biosystems UK Biobank Axiom | Illumina Infinium Global Screening Array | HumanOmni1-Quad; HumanOmni5-Quad;  Human1M-Duov3_B; HumanOmniExpress-12v1.0; HumanOmniExpress; Human660W-Quadv1_A | Illumina Global Screening Array |
| **Imputation panel** | HRC and UK10K + 1000 Genomes Phase 3 | TOPMed | TOPMed | 1000 Genomes phase 3 | Based on whole genome sequencing data from 8,429 Danes and 7,146 samples from North-West Europe |
| **Variant filters** | Imputation quality score ≥0.3; MAC_cases_ ≥10; MAC_controls_ ≥10 | | | Imputation quality score ≥0.7; MAF ≥1%. | Imputation quality score ≥0.3; MAC_cases_ ≥10; MAC_controls_ ≥10 |
| **# variants passing filters (chronic dry cough)** | EUR: 19,494,967 SAS: 6,402,724 | – | – | – | – |
| **# variants passing filters (ACEi-induced cough)** | AFR: 6,963,210 EAS: 4,460,597 EUR: 17,101,867 SAS: 8,455,429 | EUR: 6,808,289 | SAS: 9,494,374 | AFR, EUR, ‘Other’: 1,931,833 | EUR: 10,559,321 |
| **Association testing (additive genetic model)** | | | | | |
| **Software** | REGENIE | | | PLINK 1.07 | BOLT-LMM |
| **Model** | Approximate Firth logistic regression | | | Logistic regression | Linear mixed model |
| **Covariates** | Age, age², sex, genotyping array, ever smoking status (chronic dry cough only), PCs 1-10 | Age, age², sex, PCs 1-10 | Age, age², sex, PCs 1-20 | Birth year, sex, PCs 1-10 | Age, age², sex, PCs 1-10 |

Supplementary Table 2. Sample sizes and demographics.
Age for chronic dry cough is age at baseline recruitment, while age for ACEi-induced cough in UK Biobank, EXCEED and Genes & Health is age at index date (i.e.at first ACEi prescription). Abbreviations: AFR, African; EAS, East Asian; EUR, European; SAS, South Asian; SD, standard deviation.

| **Cohort** | **Ancestral group** | **Total sample size (% female)** | **Case count  (% female)** | **Control count  (% female)** | **Mean age (SD)  in total sample, years** | **Mean age (SD) in cases, years** | **Mean age (SD) in controls, years** |
| --- | --- | --- | --- | --- | --- | --- | --- |
| **Chronic dry cough** | | | | | | | |
| **UK Biobank** | EUR | 95399 (57.5) | 7575 (59.1) | 87824 (57.3) | 55.8 (7.7) | 56.8 (7.5) | 55.7 (7.7) |
|  | SAS | 889 (47.7) | 60 (60.0) | 829 (46.8) | 52.6 (8.0) | 54.1 (7.8) | 52.4 (8.0) |
| **ACEi-induced cough** | | | | | | | |
| **UK Biobank** | AFR | 408 (56.9) | 48 (75.0) | 360 (54.4) | 53.0 (8.2) | 54.5 (8.1) | 52.8 (8.2) |
|  | EAS | 148 (52.0) | 32 (46.9) | 116 (53.4) | 53.4 (7.7) | 57.5 (7.9) | 52.3 (7.2) |
|  | EUR | 37372 (42.4) | 5049 (55.0) | 32323 (40.5) | 57.9 (7.7) | 59.1 (7.2) | 57.8 (7.7) |
|  | SAS | 1101 (36.7) | 178 (43.8) | 923 (35.3) | 54.5 (8.1) | 56.3 (8.0) | 54.2 (8.0) |
| **EXCEED** | EUR | 838 (42.8) | 128 (51.6) | 710 (41.3) | 55.0 (7.2) | 55.5 (5.9) | 54.9 (7.5) |
| **Genes & Health** | SAS | 7599 (45.0) | 819 (57.8) | 6780 (43.4) | 48.5 (10.9) | 48.5 (9.9) | 48.5 (11.0) |
| **eMERGE Network (Mosley *et al,* 2016)** | Multiple ancestries (AFR, EUR, ‘other’) | 7080 (48.4) | 1595 (60.7) | 5485 (44.8) | .. | .. | .. |
| **Copenhagen Hospital Biobank** | EUR | 29546 | 13674 (..) | 15872 (..) | .. | .. | .. |

Supplementary Table 3. Independent sentinel variants identified in multi-trait GWAS of chronic dry cough and ACEi-induced cough, and additional sentinels from each single-trait GWAS.
Mapped genes in bold are novel. Direction column has the following order: UK Biobank-European, UK Biobank-South Asian for chronic dry cough; UK Biobank-African, UK Biobank-East Asian, UK Biobank-European, UK Biobank-South Asian, EXCEED-European, Genes & Health-South Asian, eMERGE Network, Copenhagen Hospital Biobank for ACEi-induced cough; chronic dry cough, ACEi-induced cough for multi-trait analysis. *Abbreviations:* CI, confidence interval; OR, odds ratio.

| **Sentinel ID** | **Position (hg19)** | **Effect  allele** | **Other  allele** | **Trait** | **Effect allele  frequency** | **OR [95% CI]** | ***p*-value** | ***N* cases** | ***N* controls** | **Direction** | **Annotation** | **Mapped gene(s)** | **Novel sentinel variant?** | **Previously reported sentinels within locus** | | |
| --- | --- | --- | --- | --- | --- | --- | --- | --- | --- | --- | --- | --- | --- | --- | --- | --- |
|  |  |  |  |  |  |  |  |  |  |  |  |  |  | **ID (*r^2^*)** | **Reported gene** | **Ref** |
| rs1544730 | 2:45588067 | A | G | Multi-trait | 0.223 | 1.11 [1.10, 1.16] | 1.72E-16 | 27515 | 145377 | ++ | Intergenic | *SRBD1* | N | rs1544730 (1.00) | *SRBD1* | ^8^ |
| 4:21393932:T:TAAG | 4:21393932 | T | TAAG | Multi-trait | 0.419 | 1.13 [1.14, 1.20] | 4.97E-32 | 27563 | 145737 | ++ | Intronic | *KCNIP4* | N | rs16870989 (0.711)  rs145489027 (0.709) | *KCNIP4*  *KCNIP4* | ^8^  ^9^ |
| 5:138404361:A:AAAG | 5:138404361 | A | AAAG | Multi-trait | 0.326 | 1.08 [1.03, 1.13] | 2.62E-08 | 12894 | 122015 | ++ | Intronic | ***CTNNA1; SIL1*** | Y | – | – | – |
| rs7761208 | 6:105776289 | T | C | Multi-trait | 0.788 | 0.89 [0.84, 0.89] | 1.72E-20 | 27531 | 145621 | -- | Intronic | *PREP* | N | rs12210271 (0.982) | *PREP* | ^8^ |
| rs7848821 | 9:128251930 | A | G | Multi-trait | 0.320 | 1.08 [1.05, 1.11] | 4.02E-12 | 29158 | 151222 | ++ | Intronic | ***MAPKAP1*** | N | rs360206 (0.158) | *SCAI* | ^8^ |
| rs112658458 | 11:49790181 | A | G | Multi-trait | 0.967 | 1.19 [1.14, 1.32] | 8.31E-09 | 27117 | 142799 | ++ | Intronic | ***OR4C12; OR4C13*** | Y | – | – | – |
| rs78598167 | 18:6110865 | T | C | Multi-trait | 0.023 | 1.33 [1.31, 1.56] | 8.36E-15 | 27295 | 143722 | ++ | Intronic | *L3MBTL4* | Y | rs8097200 (0.003) | *L3MBTL4* | ^8^ |
| rs343240 | 18:6308062 | T | C | Multi-trait | 0.178 | 0.91 [0.83, 0.89] | 4.90E-11 | 27563 | 145737 | +- | Intronic | *L3MBTL4* | N | rs8097200 (0.986) | *L3MBTL4* | ^8^ |
| rs6062847 | 20:61322018 | T | C | Multi-trait | 0.137 | 1.18 [1.15, 1.24] | 1.10E-31 | 29066 | 150277 | ++ | Intergenic | *NTSR1; SLCO4A1* | N | rs6062847 (1.00) | *NTSR1; SLCO4A1* | ^8^ |
| rs5924943 | X:150594104 | T | C | Multi-trait | 0.469 | 1.07 [1.07, 1.12] | 3.76E-16 | 27531 | 145621 | ++ | Intergenic | ***VMA21*** | Y | – | – | – |
| rs141733360 | 4:14727977 | A | C | Chronic dry cough | 0.009 | 0.51 [0.41, 0.64] | 4.20E-09 | 6254 | 41212 | -? | ncRNA intronic | ***CPEB2*** | Y | – | – | – |
| rs7518061 | 1:111087944 | T | C | ACEi-induced cough | 0.310 | 0.88 [0.84, 0.92] | 1.85E-09 | 7849 | 46697 | ---+--?? | Intergenic | ***KCNA10****;* ***RBM15*** | N | rs7526729 (0.995) | *KCNA2* | ^8^ |
| rs11172406 | 12:58366606 | A | C | ACEi-induced cough | 0.361 | 1.11 [1.07, 1.15] | 1.73E-08 | 5259 | 33362 | +-+++++? | Intergenic | ***ATP23****;* ***CYP27B1*** | Y | – | – | – |
| rs35336617 | 12:85913015 | T | TC | ACEi-induced cough | 0.703 | 0.87 [0.83, 0.91] | 2.71E-09 | 7575 | 87824 | ?---???? | Intergenic | ***ALX1****;* ***RASSF9*** | Y | – | – | – |

Supplementary Table 4. Association statistics for sentinels from multi-trait GWAS of chronic dry cough and ACEi-induced cough, in each single-trait GWAS.
Direction column has the following order: UK Biobank-European, UK Biobank-South Asian for chronic dry cough; UK Biobank-African, UK Biobank-East Asian, UK Biobank-European, UK Biobank-South Asian, EXCEED-European, Genes & Health-South Asian, eMERGE Network, Copenhagen Hospital Biobank for ACEi-induced cough. Mapped genes in bold are novel. *Abbreviations*: CI, confidence interval; OR, odds ratio.

| **ID** | **Position (hg19)** | **Effect allele** | **Other allele** | **Cough trait** | **Effect allele**  **frequency** | **OR [95% CI]** | ***p*-value** | **Direction** | **Mapped gene(s)** |
| --- | --- | --- | --- | --- | --- | --- | --- | --- | --- |
| rs1544730 | 2:45588067 | A | G | Chronic dry cough | 0.210 | 1.07 [1.02, 1.11] | 2.23E-03 | +- | *SRBD1* |
|  |  |  |  | ACEi-induced cough | 0.230 | 1.13 [1.10, 1.16] | 1.60E-15 | ?+++++?+ |  |
| 4:21393932:T:TAAG | 4:21393932 | T | TAAG | Chronic dry cough | 0.410 | 1.06 [1.03, 1.10] | 4.18E-04 | ++ | *KCNIP4* |
|  |  |  |  | ACEi-induced cough | 0.424 | 1.17 [1.14, 1.20] | 1.96E-33 | +-++++?+ |  |
| 5:138404361:A:AAAG | 5:138404361 | A | AAAG | Chronic dry cough | 0.326 | 1.09 [1.05, 1.13] | 5.99E-06 | ++ | ***CTNNA1; SIL1*** |
|  |  |  |  | ACEi-induced cough | 0.326 | 1.08 [1.03, 1.13] | 1.15E-03 | ?+++???? |  |
| rs7761208 | 6:105776289 | T | C | Chronic dry cough | 0.804 | 0.94 [0.90, 0.98] | 5.19E-03 | -- | *PREP* |
|  |  |  |  | ACEi-induced cough | 0.779 | 0.86 [0.84, 0.89] | 4.36E-21 | +?--+-?- |  |
| rs7848821 | 9:128251930 | A | G | Chronic dry cough | 0.310 | 1.07 [1.03, 1.11] | 3.81E-04 | ++ | ***MAPKAP1*** |
|  |  |  |  | ACEi-induced cough | 0.325 | 1.08 [1.05, 1.11] | 2.10E-09 | +-+-++++ |  |
| rs112658458 | 11:49790181 | A | G | Chronic dry cough | 0.965 | 1.13 [1.02, 1.24] | 1.56E-02 | +? | ***OR4C12; OR4C13*** |
|  |  |  |  | ACEi-induced cough | 0.968 | 1.23 [1.14, 1.32] | 6.30E-08 | ??+??-?+ |  |
| rs78598167 | 18:6110865 | T | C | Chronic dry cough | 0.018 | 1.13 [1.00, 1.29] | 5.66E-02 | +? | *L3MBTL4* |
|  |  |  |  | ACEi-induced cough | 0.025 | 1.43 [1.31, 1.56] | 6.10E-16 | ??++?+?+ |  |
| rs343240 | 18:6308062 | T | C | Chronic dry cough | 0.176 | 1.01 [0.97, 1.06] | 5.64E-01 | ++ | *L3MBTL4* |
|  |  |  |  | ACEi-induced cough | 0.178 | 0.86 [0.83, 0.89] | 5.18E-18 | ------?- |  |
| rs6062847 | 20:61322018 | T | C | Chronic dry cough | 0.135 | 1.16 [1.10, 1.21] | 2.07E-09 | +? | *NTSR1; SLCO4A1* |
|  |  |  |  | ACEi-induced cough | 0.138 | 1.19 [1.15, 1.24] | 4.31E-24 | -?+-++++ |  |
| rs5924943 | X:150594104 | T | C | Chronic dry cough | 0.467 | 1.03 [1.00, 1.06] | 4.40E-02 | ++ | ***VMA21*** |
|  |  |  |  | ACEi-induced cough | 0.470 | 1.09 [1.07, 1.12] | 7.96E-18 | +?++-+?+ |  |

Supplementary Table 5. Summary of protein-coding genes identified by variant-to-gene mapping (see Figure 3).
Variant positions are reported in genome build GRCh37. QTL supporting variants described are colocalising hits identified by coloc.susie.

| **Trait** | **Gene symbol** | **Ensembl ID** | **Supporting analyses** | **Supporting variants** | **Novel gene?** | **PMID** |
| --- | --- | --- | --- | --- | --- | --- |
| Multi-trait | *KCNIP4* | ENSG00000185774 | nearest; pathogenicity; PoPs | nearest(4:21393932:T:TAAG [4:21393932]); pathogenicity(4:21393932:T:TAAG [4:21393932]); PoPS(4:21393932:T:TAAG [4:21393932]) | N | 35751511; 26169577 |
| Multi-trait | *MAPKAP1* | ENSG00000119487 | nearest; pathogenicity; PoPs | nearest(rs7848821 [9:128251930]); pathogenicity(rs7046471 [9:128250409]); PoPS(rs7848821 [9:128251930]) | Y |  |
| Multi-trait | *PREP* | ENSG00000085377 | nearest; PoPs; eQTL | nearest(rs7761208 [6:105776289]); PoPS(rs7761208 [6:105776289]); eQTL(rs11156437 [6:105778067]) | N | 35751511 |
| ACEi-induced cough | *ALX1* | ENSG00000180318 | nearest; PoPs | nearest(rs35336617 [12:85913015]); PoPS(rs35336617 [12:85913015]) | Y |  |
| ACEi-induced cough | *ATP23* | ENSG00000166896 | nearest; eQTL | nearest(rs11172406 [12:58366606]); eQTL(rs4627125 [12:58372886]) | Y |  |
| Chronic dry cough | *CPEB2* | ENSG00000137449 | nearest; PoPs | nearest(rs141733360 [4:14727977]); PoPS(rs141733360 [4:14727977]) | Y |  |
| Multi-trait | *L3MBTL4* | ENSG00000154655 | nearest; PoPs | nearest(rs78598167 [18:6110865]); nearest(rs343240 [18:6308062]); PoPS(rs78598167 [18:6110865]); PoPS(rs343240 [18:6308062]) | N | 35751511 |
| Multi-trait | *OR4C13* | ENSG00000258817 | nearest; pathogenicity | nearest(rs112658458 [11:49790181]); pathogenicity(rs112658458 [11:49790181]) | Y |  |
| Multi-trait | *SIL1* | ENSG00000120725 | nearest; eQTL | nearest(5:138404361:A:AAAG [5:138404361]); eQTL(rs12173085 [5:138480025]); eQTL(rs55650316 [5:138467292]) | Y |  |
| Multi-trait | *SRBD1* | ENSG00000068784 | nearest; PoPs | nearest(rs1544730 [2:45588067]); PoPS(rs1544730 [2:45588067]) | N | 35751511 |
| Multi-trait | *CTNNA1* | ENSG00000044115 | PoPs | PoPS(5:138404361:A:AAAG [5:138404361]) | Y |  |
| ACEi-induced cough | *CYP27B1* | ENSG00000111012 | PoPs | PoPS(rs11172406 [12:58366606]) | Y |  |
| ACEi-induced cough | *KCNA10* | ENSG00000143105 | nearest | nearest(rs7518061 [1:111087944]) | Y |  |
| Multi-trait | *NTSR1* | ENSG00000101188 | nearest | nearest(rs6062847 [20:61322018]) | N | 35751511 |
| Multi-trait | *OR4C12* | ENSG00000221954 | PoPs | PoPS(rs112658458 [11:49790181]) | Y |  |
| ACEi-induced cough | *RASSF9* | ENSG00000198774 | eQTL | eQTL(rs2199514 [12:85991536]) | Y |  |
| ACEi-induced cough | *RBM15* | ENSG00000162775 | PoPs | PoPS(rs7518061 [1:111087944]) | Y |  |
| Multi-trait | *SLCO4A1* | ENSG00000101187 | PoPs | PoPS(rs6062847 [20:61322018]) | N | 35751511 |
| Multi-trait | *VMA21* | ENSG00000160131 | nearest | nearest(rs5924943 [23:150594104]) | Y |  |

#### **Supplementary Table 6. Variants with posterior inclusion probability (PIP) ≥10% identified through fine-mapping of multi-trait loci and additional independent loci from each single-trait GWAS.** *Abbreviations*: CI, confidence interval; OR, odds ratio.

| **Trait** | **Locus sentinel** | **Variant** | **Variant position  (hg19)** | **Effect allele** | **Other allele** | **Effect allele frequency** | **Posterior inclusion  probability** | **OR [95% CI]** | ***p*-value** | **Nearest gene** | **Annotation** | **CADD score** |
| --- | --- | --- | --- | --- | --- | --- | --- | --- | --- | --- | --- | --- |
| Multi-trait | rs1544730 | rs1544730 | 2:45588067 | A | G | 0.223 | 0.375 | 1.11 [1.08, 1.13] | 1.72E-16 | *SRBD1* | Intergenic | 5.231 |
| Multi-trait | rs1544730 | rs2112037 | 2:45591129 | T | C | 0.222 | 0.140 | 1.11 [1.08, 1.13] | 5.36E-16 | *SRBD1* | Intergenic | 0.854 |
| Multi-trait | rs1544730 | rs3755070 | 2:45629316 | T | C | 0.230 | 0.131 | 1.1 [1.08, 1.13] | 5.93E-16 | *SRBD1* | Intronic | 5.647 |
| Multi-trait | 4:21393932:T:TAAG | 4:21393932:T:TAAG | 4:21393932 | T | TAAG | 0.419 | 1.000 | 1.13 [1.11, 1.15] | 4.97E-32 | *KCNIP4* | Intronic | 17.44 |
| Multi-trait | 5:138404361:A:AAAG | 5:138404361:A:AAAG | 5:138404361 | A | AAAG | 0.326 | 0.140 | 1.08 [1.05, 1.11] | 2.62E-08 | *SIL1* | Intronic | 0.709 |
| Multi-trait | rs7848821 | rs7848821 | 9:128251930 | A | G | 0.320 | 0.393 | 1.08 [1.05, 1.10] | 4.02E-12 | *MAPKAP1* | Intronic | 8.129 |
| Multi-trait | rs7848821 | rs7046471 | 9:128250409 | T | C | 0.685 | 0.346 | 0.93 [0.91, 0.95] | 4.61E-12 | *MAPKAP1* | Intronic | 14 |
| Multi-trait | rs7848821 | rs11794506 | 9:128251275 | A | G | 0.323 | 0.133 | 1.08 [1.05, 1.10] | 1.34E-11 | *MAPKAP1* | Intronic | 5.953 |
| Multi-trait | rs112658458 | rs112658458 | 11:49790181 | A | G | 0.967 | 0.189 | 1.19 [1.12, 1.26] | 8.31E-09 | *OR4C13* | Intronic | 13.18 |
| Multi-trait | rs112658458 | rs145450510 | 11:49733467 | T | C | 0.033 | 0.170 | 0.84 [0.79, 0.89] | 9.43E-09 | *OR4C13* | Intronic | 2.287 |
| Multi-trait | rs112658458 | rs72914105 | 11:49643260 | T | C | 0.967 | 0.140 | 1.19 [1.12, 1.26] | 1.20E-08 | *OR4C13* | Intronic | 4.918 |
| Multi-trait | rs112658458 | rs182343842 | 11:49861001 | T | C | 0.032 | 0.140 | 0.84 [0.79, 0.89] | 1.17E-08 | *OR4C13* | Intergenic | 1.105 |
| Multi-trait | rs112658458 | rs72914160 | 11:49716385 | T | C | 0.967 | 0.110 | 1.19 [1.12, 1.26] | 1.43E-08 | *OR4C13* | Intronic | 1.953 |
| Multi-trait | rs78598167 | rs78598167 | 18:6110865 | T | C | 0.023 | 0.988 | 1.32 [1.23, 1.42] | 8.36E-15 | *L3MBTL4* | Intronic | 1.213 |
| Multi-trait | rs6062847 | rs6062847 | 20:61322018 | T | C | 0.137 | 1.000 | 1.18 [1.15, 1.22] | 1.10E-31 | *NTSR1* | Intergenic | 3.435 |
| Multi-trait | rs5924943 | rs5924943 | 23:150594104 | T | C | 0.469 | 0.292 | 1.07 [1.05, 1.09] | 3.76E-16 | *VMA21* | Intergenic | 1.199 |
| Multi-trait | rs5924943 | rs12007715 | 23:150597999 | A | C | 0.472 | 0.113 | 1.07 [1.05, 1.09] | 9.88E-16 | *VMA21* | Intergenic | 8.138 |
| Chronic dry cough | rs141733360 | rs141733360 | 4:14727977 | A | C | 0.009 | 0.997 | 0.51 [0.41, 0.64] | 4.20E-09 | *CPEB2* | ncRNA intronic | 1.376 |
| ACEi-induced cough | rs7518061 | rs7518061 | 1:111087944 | T | C | 0.310 | 0.453 | 0.88 [0.84, 0.92] | 1.85E-09 | *KCNA10* | Intergenic | 0.733 |
| ACEi-induced cough | rs7518061 | rs7526729 | 1:111087058 | A | G | 0.688 | 0.440 | 1.14 [1.09, 1.19] | 2.16E-09 | *KCNA10* | Intergenic | 5.952 |
| ACEi-induced cough | rs11172406 | rs11172406 | 12:58366606 | A | C | 0.361 | 0.605 | 1.11 [1.07, 1.15] | 1.73E-08 | *ATP23* | Intergenic | 3.193 |
| ACEi-induced cough | rs11172406 | rs6581173 | 12:58415737 | T | C | 0.365 | 0.141 | 1.11 [1.07, 1.15] | 7.83E-08 | *ATP23* | Intergenic | 3.683 |
| ACEi-induced cough | rs35336617 | rs35336617 | 12:85913015 | T | TC | 0.703 | 0.328 | 0.87 [0.83, 0.91] | 2.71E-09 | *ALX1* | Intergenic | 0.5 |
| ACEi-induced cough | rs35336617 | 12:85838772_G_GA | 12:85838772 | G | GA | 0.306 | 0.133 | 1.13 [1.09, 1.18] | 6.85E-09 | *ALX1* | Intergenic | 0.099 |
| ACEi-induced cough | rs35336617 | rs548204060 | 12:86033240 | CA | C | 0.375 | 0.111 | 1.14 [1.09, 1.19] | 8.02E-09 | *RASSF9* | Intergenic | 1.145 |

#### **Supplementary Table 7. Genes with highest polygenic priority score (PoPs) within specified window of sentinels identified in the multi-trait GWAS and additional sentinels variants from each single-trait GWAS.**

| **Trait** | **Sentinel** | **Position (hg19)** | **Gene symbol** | **Ensembl ID** | **Polygenic priority score** | **Window (bp)** |
| --- | --- | --- | --- | --- | --- | --- |
| Multi-trait | rs1544730 | 2:45588067 | *SRBD1* | ENSG00000068784 | 0.5313 | 250000 |
| Multi-trait | 4:21393932:T:TAAG | 4:21393932 | *KCNIP4* | ENSG00000185774 | 0.7403 | 250000 |
| Multi-trait | 5:38404361:A:AAAG | 5:138404361 | *CTNNA1* | ENSG00000044115 | 0.4941 | 250000 |
| Multi-trait | rs7761208 | 6:105776289 | *PREP* | ENSG00000085377 | 0.5370 | 250000 |
| Multi-trait | rs7848821 | 9:128251930 | *MAPKAP1* | ENSG00000119487 | 0.6797 | 250000 |
| Multi-trait | rs112658458 | 11:49790181 | *OR4C12* | ENSG00000221954 | 0.0051 | 250000 |
| Multi-trait | rs78598167 | 18:6110865 | *L3MBTL4* | ENSG00000154655 | 0.7104 | 250000 |
| Multi-trait | rs343240 | 18:6308062 | *L3MBTL4* | ENSG00000154655 | 0.7104 | 250000 |
| Multi-trait | rs6062847 | 20:61322018 | *SLCO4A1* | ENSG00000101187 | 0.3588 | 250000 |
| Chronic dry cough | rs141733360 | 4:14727977 | *CPEB2* | ENSG00000137449 | -0.1807 | 500000 |
| ACEi-induced cough | rs7518061 | 1:111087944 | *RBM15* | ENSG00000162775 | 0.4918 | 250000 |
| ACEi-induced cough | rs11172406 | 12:58366606 | *CYP27B1* | ENSG00000111012 | 0.7569 | 250000 |
| ACEi-induced cough | rs35336617 | 12:85913015 | *ALX1* | ENSG00000180318 | -0.1156 | 250000 |

Supplementary Table 8. QTL colocalisation results.
Results are filtered for evidence for colocalisation between GWAS and expression/protein QTLs (H4) ≥0.7.

| **QTL** | **Trait** | **Locus sentinel** | **Locus sentinel position**  **(hg19)** | **Dataset** | **Tissue** | **Gene** | **Ensembl ID** | **hit1** | **hit2** | **H0** | **H1** | **H2** | **H3** | **H4** |
| --- | --- | --- | --- | --- | --- | --- | --- | --- | --- | --- | --- | --- | --- | --- |
| eQTL | Multi-trait | rs11949916 | 5:138396277 | GTeX_v8 | Lung | *SIL1* | ENSG00000120725 | 5_138467292_C_G | 5_138313146_A_G | 5.86E-09 | 2.88E-06 | 5.56E-04 | 2.72E-01 | 7.28E-01 |
| eQTL | Multi-trait | rs11949916 | 5:138396277 | eQTLGen | Whole_Blood | *SIL1* | ENSG00000120725 | 5_138480025_A_G | 5_138377550_C_G | 0.00E+00 | 0.00E+00 | 2.67E-04 | 1.24E-01 | 8.76E-01 |
| eQTL | Multi-trait | rs11949916 | 5:138396277 | eQTLGen | Whole_Blood | *SIL1* | ENSG00000120725 | 5_138480025_A_G | 5_138392482_A_G | 0.00E+00 | 0.00E+00 | 3.48E-04 | 1.62E-01 | 8.38E-01 |
| eQTL | Multi-trait | rs11949916 | 5:138396277 | eQTLGen | Whole_Blood | *SIL1* | ENSG00000120725 | 5_138480025_A_G | 5_138398172_A_G | 0.00E+00 | 0.00E+00 | 3.61E-04 | 1.68E-01 | 8.32E-01 |
| eQTL | Multi-trait | rs11949916 | 5:138396277 | eQTLGen | Whole_Blood | *SIL1* | ENSG00000120725 | 5_138480025_A_G | 5_138396277_G_T | 0.00E+00 | 0.00E+00 | 2.19E-04 | 1.01E-01 | 8.99E-01 |
| eQTL | Multi-trait | rs7761208 | 6:105776289 | eQTLGen | Whole_Blood | *PREP* | ENSG00000085377 | 6_105778067_C_T | 6_105799455_C_T | 2.56E-25 | 1.54E-13 | 6.26E-14 | 3.57E-02 | 9.64E-01 |
| eQTL | ACEi-induced cough | rs11172406 | 12:58366606 | GTeX_v8 | Whole_Blood | *ATP23* | ENSG00000166896 | 12_58372886_C_T | 12_58458032_A_G | 8.11E-15 | 2.51E-13 | 7.61E-03 | 2.34E-01 | 7.59E-01 |
| eQTL | ACEi-induced cough | rs12230791 | 12:85919981 | GTeX_v8 | Adrenal_Gland | *RASSF9* | ENSG00000198774 | 12_85991536_A_G | 12_86016913_A_G | 5.40E-10 | 3.97E-06 | 1.59E-05 | 1.15E-01 | 8.85E-01 |

#### **Supplementary Table 9. Druggability.**

| **Gene** | **Drug name** | **Indication (phase)** | **ChEMBL ID** | **Experimental Factor Ontology Terms** | **Interaction type** | **Drug description** | **Approval Year** |
| --- | --- | --- | --- | --- | --- | --- | --- |
| *KCNA10* | DALFAMPRIDINE | Trauma, Nervous System(2); Guillain-Barre Syndrome(2); Optic Neuritis(0); Muscle Spasticity(3); Multiple Sclerosis, Relapsing-Remitting(3); Cerebral Palsy(1); Multiple Sclerosis(4); Motor Neuron Disease(1); Stroke(3); Muscular Atrophy, Spinal(2); Spinal Cord Injuries(3); Renal Insufficiency(1); Multiple Sclerosis, Chronic Progressive(3); Ischemic Stroke(3); Sleep Apnea, Obstructive(2) | CHEMBL284348 | nervous system injury; Guillain-Barre syndrome; optic neuritis; Spasticity; relapsing-remitting multiple sclerosis; cerebral palsy; multiple sclerosis; motor neuron disease; stroke; Proximal spinal muscular atrophy type 3; Spinal cord injury; Renal insufficiency; secondary progressive multiple sclerosis; Ischemic stroke; obstructive sleep apnea | blocker | Small molecule potassium channel blocker used to improve motor function. | 2010 |
| *KCNA10* | NERISPIRDINE | Multiple Sclerosis(2); Spinal Cord Injuries(2) | CHEMBL2107762 | multiple sclerosis; Spinal cord injury | blocker |  | NA |
| *NTSR1* | REMINERTANT | Small Cell Lung Carcinoma(2) | CHEMBL506981 | small cell lung carcinoma | antagonist |  | NA |
| *KCNA10* | TEDISAMIL | Arrhythmias, Cardiac(0); Atrial Fibrillation(3); Atrial Flutter(3) | CHEMBL2111110 | cardiac arrhythmia; atrial fibrillation; atrial flutter | blocker |  | NA |

Supplementary Table 10. Sentinel variant PheWAS results (false discovery rate [FDR] <0.01) using DeepPheWAS.
Mapped genes in bold are novel. ^*^The chronic dry cough phenotype denoted here has the same definition as ‘chronic dry cough’ described above and utilised in our genetic analyses (Appendix pp 2). A*bbreviations*: EA, effect allele; MA, minor allele; MAF, minor allele frequency; OA, other allele; OR, odds ratio; SE, standard error.

| **Trait** | **Sentinel** | **Position (hg19)** | **EA** | **OA** | **MA** | **MAF** | **Trait description** | **Ancestry** | **N** | **FDR** | ***p*-value** | **OR** | **Beta** | **L95** | **U95** | **SE** | **Direction** | **Mapped gene(s)** |
| --- | --- | --- | --- | --- | --- | --- | --- | --- | --- | --- | --- | --- | --- | --- | --- | --- | --- | --- |
| ACEi-induced cough | rs11172406 | 12:58366606 | A | C | A | 0.337 | Urate | EUR | 376291 | 7.85E-14 | 4.05E-17 | NA | -0.017 | -0.021 | -0.013 | 0.002 | - | ***ATP23****;* ***CYP27B1*** |
| Multi-trait | rs6062847 | 20:61322018 | T | C | T | 0.136 | Chronic dry cough^*^ | EUR | 96223 | 1.12E-07 | 5.76E-11 | 1.163 | NA | 1.111 | 1.216 | 0.023 | + | *NTSR1; SLCO4A1* |
| Multi-trait | rs5924943 | X:150594104 | T | C | T | 0.467 | IGF-1 | EUR | 374381 | 3.17E-06 | 1.64E-09 | NA | -0.011 | -0.015 | -0.008 | 0.002 | - | ***VMA21*** |
| Multi-trait | rs6062847 | 20:61322018 | T | C | T | 0.136 | Cough on most days | EUR | 105125 | 4.24E-06 | 4.37E-09 | 1.111 | NA | 1.072 | 1.150 | 0.018 | + | *NTSR1; SLCO4A1* |
| Chronic dry cough | rs141733360 | 4:14727977 | C | A | A | 0.009 | Chronic dry cough^*^ | EUR | 96223 | 1.98E-04 | 1.02E-07 | 1.894 | NA | 1.497 | 2.397 | 0.120 | + | ***CPEB2*** |
| ACEi-induced cough | rs11172406 | 12:58366606 | A | C | A | 0.337 | Sitting height | EUR | 392979 | 5.17E-04 | 5.34E-07 | NA | -0.009 | -0.013 | -0.005 | 0.002 | - | ***ATP23****;* ***CYP27B1*** |
| Chronic dry cough | rs141733360 | 4:14727977 | C | A | A | 0.009 | Cough on most days | EUR | 105125 | 2.18E-03 | 2.24E-06 | 1.469 | NA | 1.253 | 1.723 | 0.081 | + | ***CPEB2*** |
| ACEi-induced cough | rs11172406 | 12:58366606 | A | C | A | 0.337 | Urea | EUR | 376503 | 6.21E-03 | 9.60E-06 | NA | -0.010 | -0.015 | -0.006 | 0.002 | - | ***ATP23****;* ***CYP27B1*** |
| Multi-trait | rs343240 | 18:6308062 | C | T | T | 0.194 | Other specified cardiac dysrhythmias | SAS | 7854 | 7.74E-03 | 7.96E-06 | 0.578 | NA | 0.454 | 0.735 | 0.123 | - | *L3MBTL4* |

#### **Supplementary Table 11. Open Targets Genetics ‘GWAS lead variants’ results.** Mapped genes in bold are novel. *Abbreviations*: CI, confidence interval; OR, odds ratio; SE, standard error.

| **Trait** | **Sentinel variant** | **Query variant** | **Open Targets lead variant** | **Open Targets lead variant  tested allele** | ***r^2^*** | **Study ID** | **Study Trait** | ***p*-value** | **Beta** | **OR** | **95% CI** | **Author, Year (PMID)** | **Study N** | **Direction** | **Mapped gene(s)** |
| --- | --- | --- | --- | --- | --- | --- | --- | --- | --- | --- | --- | --- | --- | --- | --- |
| Multi-trait | 4:21393932:T:TAAG | rs7675300 | rs1495509 | C | 0.997 | GCST003027 | Cough in response to angiotensin-converting enzyme inhibitor drugs | 2.00E-09 |  | 1.23 | (1.1, 1.3) | Mosley JD, 2016 (26169577) | 12311 | + | *KCNIP4* |
| Multi-trait | 5:138404361:A:AAAG | rs11949916 | rs11242445 | T | 0.864 | GCST90013473 | Biological sex | 5.00E-10 | -0.0117 |  | (−0.015, −0.0080) | Pirastu N, 2021 (33888908) | 2462132 | - | ***CTNNA1*; *SIL1*** |
| Multi-trait | rs6062847 | rs6062847 | rs6062847 | T | 1.000 | NEALE2_22502 | Cough on most days | 1.38E-09 |  | 1.13 | (1.1, 1.2) | UKB Neale v2 | 91787 | + | *NTSR1; SLCO4A1* |
| ACEi-induced cough | rs11172406 | rs11172406 | rs12578279 | G | 0.877 | GCST006569 | Self-reported math ability (MTAG) [MTAG] | 5.00E-19 |  |  |  | Lee JJ, 2018 (30038396) | 670471 | NA | ***ATP23****;* ***CYP27B1*** |
| ACEi-induced cough | rs11172406 | rs11172406 | rs12578279 | G | 0.877 | GCST006573 | Self-reported math ability | 1.00E-16 |  |  |  | Lee JJ, 2018 (30038396) | 564698 | NA | ***ATP23****;* ***CYP27B1*** |
| ACEi-induced cough | rs11172406 | rs11172406 | rs4760364 | G | 0.877 | GCST006568 | Highest math class taken (MTAG) [MTAG] | 6.00E-18 | -0.0152 |  | (0.012, 0.019) | Lee JJ, 2018 (30038396) | 811539 | - | ***ATP23****;* ***CYP27B1*** |
| ACEi-induced cough | rs11172406 | rs11172406 | rs10877067 | T | 0.877 | GCST90025965 | Urate levels | 2.60E-29 | -0.0224 |  | (0.018, 0.026) | Barton AR, 2021 (34226706) | 437354 | - | ***ATP23****;* ***CYP27B1*** |

#### **Supplementary Table 12. Description of studies included in Respiratory PheWAS.**

| **Clinical phenotype** | **Clinical subtype** | **Cohort** | **Ancestry** | ***N*** | **Author (Year)** | **Reference** | **Additional information** |
| --- | --- | --- | --- | --- | --- | --- | --- |
| Asthma | Asthma | GBMI | MULTI | 1800785 | Tsuo (2022) | ^31^ |  |
| Asthma | Asthma | UK Biobank; deCODE | EUR | 771388 | Olafsdottir (2020) | ^32^ |  |
| Asthma | Asthma adult onset | UK Biobank | EUR | 327253 | Ferreira (2019) | ^33^ |  |
| Asthma | Asthma childhood onset | UK Biobank | EUR | 314633 | Ferreira (2019) | ^33^ |  |
| Asthma | Asthma exacerbations in children | Danish | EUR | 68281 | Ahluwalia (2020) | ^34^ |  |
| Asthma | Asthma exacerbations in adults | UK Biobank | EUR | 49494 | Edris (2024) | ^35^ |  |
| Bronchiectasis | Bronchiectasis | UK Biobank; FinnGen;Biobank Japan | MULTI | 605195 | Sakaue (2021) | ^36^ |  |
| Bronchopneumonia | Bronchopneumonia | UK Biobank | EUR | 456348 | Jiang (2021) | ^37^ |  |
| Bronchopneumonia | Bronchopneumonia | UK Biobank | EUR | 387930 | Backman (2021) | ^38^ |  |
| Chronic bronchitis | Chronic bronchitis | UK Biobank | EUR | 378459 | DeepPheWAS | ^11^ | Defined in UK Biobank using DeepPheWAS |
| Chronic bronchitis | Chronic bronchitis | FinnGen | EUR | 339565 | FinnGen | ^39^ | FinnGen DF10: J10_BRONCHNAS |
| Chronic obstructive pulmonary disease | Chronic obstructive pulmonary disease | UK Biobank | EUR | 196578 | DeepPheWAS | ^11^ | Defined in UK Biobank using DeepPheWAS |
| Chronic obstructive pulmonary disease | Chronic obstructive pulmonary disease | FinnGen | EUR | 358369 | FinnGen | ^39^ | FinnGen DF10: J10_COPD |
| Chronic obstructive pulmonary disease | Chronic obstructive pulmonary disease with acute exacerbation | UK Biobank;GSK | EUR | 50365 | Williams | NA |  |
| Chronic sputum production | Chronic sputum production | UK Biobank | EUR | 58185 | Packer (2023) | ^40^ |  |
| Emphysema | Emphysema | UK Biobank | EUR | 178144 | DeepPheWAS | ^11^ | Defined in UK Biobank using DeepPheWAS |
| Emphysema | Emphysema | FinnGen | EUR | 340431 | FinnGen | ^39^ | FinnGen DF10: J10_EMPHYSEMA |
| Idiopathic Pulmonary Fibrosis | Idiopathic pulmonary fibrosis | GBMI | MULTI | 1375570 | Partanen (2022) | ^41^ |  |
| Idiopathic Pulmonary Fibrosis | Idiopathic pulmonary fibrosis | US;UK;Spain | EUR | 24589 | Allen (2022) | ^42^ |  |
| Idiopathic Pulmonary Fibrosis | Idiopathic pulmonary fibrosis DCLO | US;UK;Spain | EUR | 975 | Allen (2023) | ^43^ |  |
| Idiopathic Pulmonary Fibrosis | Idiopathic pulmonary fibrosis FVC | US;UK;Spain | EUR | 1329 | Allen (2023) | ^43^ |  |
| Idiopathic Pulmonary Fibrosis | Idiopathic pulmonary fibrosis Survival | US;UK;Spain | EUR | 1481 | Oldham (2023) | ^44^ |  |
| Respiratory infections | Respiratory infections primary care | UK Biobank | EUR | 40047 | Williams | NA |  |
| Respiratory infections | Respiratory infections secondary care | UK Biobank | EUR | 120897 | Williams (2023) | ^45^ |  |
| Interstitial lung abnormality | Interstitial lung abnormality | FHS;AGES;COPDGene;ECLIPSE;MESA;SPIROMICS | MULTI | 11973 | Hobbs (2019) | ^46^ |  |
| Interstitial lung abnormality | Subpleural predominant interstitial lung abnormality | FHS;AGES;COPDGene;ECLIPSE;MESA;SPIROMICS | MULTI | 11561 | Hobbs (2019) | ^46^ |  |

Supplementary Table 13. Sentinel variant Respiratory PheWAS results (*p*-value <0.001).
Mapped genes in bold are novel. *Abbreviations*: EA, effect allele; OA, other allele OR, odds ratio; SE, standard error.

| **Trait** | **Sentinel** | **Position (hg19)** | **EA** | **OA** | **Clinical phenotype** | **Clinical subtype** | **Cohort** | **Ancestry** | **Author (Year)** | **PMID** | ***p*-value** | **N** | **Beta** | **OR** | **L95** | **U95** | **SE** | **Direction** | **Mapped gene(s)** |
| --- | --- | --- | --- | --- | --- | --- | --- | --- | --- | --- | --- | --- | --- | --- | --- | --- | --- | --- | --- |
| Multi-trait | rs6062847 | 20:61322018 | T | C | Asthma | Asthma | GBMI | Multi | Tsuo (2022) | 36778051 | 1.70E-05 | 1800785 | 0.0261 | 1.03 | 1.01 | 1.04 | 0.0061 | + | *NTSR1; SLCO4A1* |
| Multi-trait | rs7848821 | 9:128251930 | A | G | Asthma | Asthma | GBMI | Multi | Tsuo (2022) | 36778051 | 3.70E-04 | 1800785 | 0.0158 | 1.02 | 1.01 | 1.02 | 0.0044 | + | ***MAPKAP1*** |

Supplementary Table 14. UK Biobank asthma exclusions sensitivity analysis.
Mapped genes in bold are novel. In the primary analysis, direction column has the following order: UK Biobank-European, UK Biobank-South Asian for chronic dry cough; UK Biobank-African, UK Biobank-East Asian, UK Biobank-European, UK Biobank-South Asian, EXCEED-European, Genes & Health-South Asian, eMERGE Network, Copenhagen Hospital Biobank for ACEi-induced cough; chronic dry cough, ACEi-induced cough for multi-trait analysis. In the UK Biobank only (excluding asthma) sensitivity analysis, direction column has the following order: UK Biobank-European, UK Biobank-South Asian for chronic dry cough; UK Biobank-African, UK Biobank-East Asian, UK Biobank-European, UK Biobank-South Asian; chronic dry cough, ACEi-induced cough for multi-trait analysis. *Abbreviations*: EAF, effect allele frequency; CI, confidence interval; OR, odds ratio.

| **Trait** | **Sentinel** | **Position (hg19)** | **Effect Allele** | **Other Allele** | **Primary analysis** | | | | | | **UK Biobank only (excluding asthma)** | | | | | | **Mapped gene(s)** |
| --- | --- | --- | --- | --- | --- | --- | --- | --- | --- | --- | --- | --- | --- | --- | --- | --- | --- |
|  |  |  |  |  | **EAF** | **OR [95% CI]** | ***p*-value** | ***N*  cases** | ***N* controls** | **Direction** | **EAF** | **OR [95% CI]** | ***p*-value** | ***N*  cases** | ***N* controls** | **Direction** |  |
| Multi-trait | rs1544730 | 2:45588067 | A | G | 0.223 | 1.11 [1.08, 1.13] | 1.72E-16 | 27515 | 145377 | ++ | 0.210 | 1.09 [1.06, 1.13] | 5.98E-07 | 10539 | 110137 | ++ | *SRBD1* |
| Multi-trait | 4:21393932:T:TAAG | 4:21393932 | T | TAAG | 0.419 | 1.13 [1.11, 1.15] | 4.97E-32 | 27563 | 145737 | ++ | 0.411 | 1.13 [1.09, 1.16] | 2.07E-15 | 10584 | 110442 | ++ | *KCNIP4* |
| Multi-trait | 5:138404361:A:AAAG | 5:138404361 | A | AAAG | 0.326 | 1.08 [1.05, 1.11] | 2.62E-08 | 12894 | 122015 | ++ | 0.326 | 1.07 [1.04, 1.11] | 6.16E-06 | 10539 | 110137 | ++ | ***CTNNA1; SIL1*** |
| Multi-trait | rs7761208 | 6:105776289 | T | C | 0.788 | 0.89 [0.87, 0.91] | 1.72E-20 | 27531 | 145621 | -- | 0.802 | 0.88 [0.85, 0.91] | 4.14E-12 | 10555 | 110339 | -- | *PREP* |
| Multi-trait | rs7848821 | 9:128251930 | A | G | 0.320 | 1.08 [1.05, 1.10] | 4.02E-12 | 29158 | 151222 | ++ | 0.311 | 1.09 [1.06, 1.12] | 6.28E-08 | 10584 | 110442 | ++ | ***MAPKAP1*** |
| Multi-trait | rs112658458 | 11:49790181 | A | G | 0.967 | 1.19 [1.12, 1.26] | 8.31E-09 | 27117 | 142799 | ++ | 0.966 | 1.18 [1.09, 1.28] | 1.12E-04 | 10326 | 108517 | ++ | ***OR4C12; OR4C13*** |
| Multi-trait | rs78598167 | 18:6110865 | T | C | 0.023 | 1.33 [1.24, 1.43] | 8.36E-15 | 27295 | 143722 | ++ | 0.019 | 1.43 [1.29, 1.59] | 1.34E-11 | 10464 | 109278 | ++ | *L3MBTL4* |
| Multi-trait | rs343240 | 18:6308062 | T | C | 0.178 | 0.91 [0.89, 0.94] | 4.90E-11 | 27563 | 145737 | +- | 0.178 | 0.91 [0.87, 0.94] | 1.24E-06 | 10584 | 110442 | +- | *L3MBTL4* |
| Multi-trait | rs6062847 | 20:61322018 | T | C | 0.137 | 1.18 [1.15, 1.22] | 1.10E-31 | 29066 | 150277 | ++ | 0.135 | 1.20 [1.15, 1.25] | 1.80E-18 | 10509 | 109583 | ++ | *NTSR1; SLCO4A1* |
| Multi-trait | rs5924943 | X:150594104 | T | C | 0.469 | 1.07 [1.05, 1.09] | 3.76E-16 | 27531 | 145621 | ++ | 0.468 | 1.06 [1.04, 1.09] | 1.03E-06 | 10555 | 110339 | ++ | ***VMA21*** |
| Chronic dry cough | rs141733360 | 4:14727977 | A | C | 0.009 | 0.51 [0.41, 0.64] | 4.20E-09 | 7575 | 87824 | -? | 0.009 | 0.58 [0.46, 0.74] | 1.05E-05 | 6202 | 81172 | -? | ***CPEB2*** |
| ACEi-induced cough | rs7518061 | 1:111087944 | T | C | 0.310 | 0.88 [0.84, 0.92] | 1.85E-09 | 6254 | 41212 | ---+--?? | 0.304 | 0.87 [0.83, 0.92] | 2.86E-07 | 4336 | 28514 | ---+ | ***KCNA10; RBM15*** |
| ACEi-induced cough | rs11172406 | 12:58366606 | A | C | 0.361 | 1.11 [1.07, 1.15] | 1.73E-08 | 7849 | 46697 | +-+++++? | 0.340 | 1.12 [1.07, 1.18] | 2.80E-06 | 4336 | 28514 | +-++ | ***ATP23; CYP27B1*** |
| ACEi-induced cough | rs35336617 | 12:85913015 | T | TC | 0.703 | 0.87 [0.83, 0.91] | 2.71E-09 | 5259 | 33362 | ?---???? | 0.704 | 0.87 [0.82, 0.91] | 4.24E-08 | 4291 | 28209 | ?--+ | ***ALX1; RASSF9*** |

Supplementary Table 15. Genetic risk score PheWAS results (false discovery rate [FDR] <0.01) using DeepPheWAS.
*Abbreviations*: OR, odds ratio; SE, standard error.

| **PheWAS ID** | **Phenotype** | **FDR** | ***p*-value** | **OR** | **Beta** | **L95** | **U95** | **SE** | **Phenotype group** | **Phenotype group (narrow)** | **Direction** |
| --- | --- | --- | --- | --- | --- | --- | --- | --- | --- | --- | --- |
| Q0526 | Testosterone | 3.18E-302 | 1.93E-294 | NA | -0.32 | -0.33 | -0.3 | 0.0084 | Quantitative Measure | Blood Biochemistry | - |
| Q0008 | Whole body fat-free mass | 1.23E-286 | 1.27E-289 | NA | -0.29 | -0.3 | -0.27 | 0.0079 | Quantitative Measure | Anthropometry | - |
| Q0012 | Hand grip strength | 5.85E-277 | 9.05E-280 | NA | -3.2 | -3.3 | -3 | 0.089 | Quantitative Measure | Body Measurements | - |
| Q0010 | Basal metabolic rate | 6.30E-269 | 1.30E-271 | NA | -0.28 | -0.29 | -0.26 | 0.0079 | Quantitative Measure | Anthropometry | - |
| Q0001 | Standing height | 7.52E-232 | 1.94E-234 | NA | -0.26 | -0.27 | -0.24 | 0.0079 | Quantitative Measure | Anthropometry | - |
| Q0007 | Waist to hip ratio | 1.56E-225 | 4.83E-228 | NA | -0.25 | -0.27 | -0.24 | 0.0079 | Quantitative Measure | Anthropometry | - |
| Q0511 | Creatinine | 1.32E-198 | 4.78E-201 | NA | -0.24 | -0.26 | -0.23 | 0.008 | Quantitative Measure | Blood Biochemistry | - |
| Q0002 | Sitting height | 9.48E-195 | 3.91E-197 | NA | -0.24 | -0.25 | -0.22 | 0.0078 | Quantitative Measure | Anthropometry | - |
| Q0603 | Haemoglobin concentration | 9.05E-165 | 4.20E-167 | NA | -0.22 | -0.24 | -0.2 | 0.008 | Quantitative Measure | Blood Count | - |
| Q0529 | Urate | 3.16E-164 | 1.63E-166 | NA | -0.22 | -0.24 | -0.21 | 0.008 | Quantitative Measure | Blood Biochemistry | - |
| Q0604 | Haematocrit percentage | 5.29E-148 | 3.00E-150 | NA | -0.21 | -0.22 | -0.19 | 0.008 | Quantitative Measure | Blood Count | - |
| Q0120 | Forced vital capacity (best) | 4.83E-146 | 2.99E-148 | NA | -0.23 | -0.25 | -0.22 | 0.009 | Quantitative Measure | Spirometry | - |
| Q0122 | Peak expiratory flow | 7.10E-141 | 4.76E-143 | NA | -0.21 | -0.23 | -0.19 | 0.0082 | Quantitative Measure | Spirometry | - |
| Q0602 | Red blood cell (erythrocyte) count | 1.12E-119 | 8.06E-122 | NA | -0.19 | -0.2 | -0.17 | 0.008 | Quantitative Measure | Blood Count | - |
| Q0121 | Forced expiratory volume in 1-second (FEV1), Best measure | 2.60E-118 | 2.01E-120 | NA | -0.21 | -0.23 | -0.19 | 0.009 | Quantitative Measure | Spirometry | - |
| Q0005 | Waist circumference | 1.07E-111 | 8.87E-114 | NA | -0.18 | -0.19 | -0.16 | 0.0079 | Quantitative Measure | Anthropometry | - |
| Q0003 | Weight | 3.22E-109 | 2.82E-111 | NA | -0.18 | -0.19 | -0.16 | 0.0079 | Quantitative Measure | Anthropometry | - |
| Q0517 | HDL cholesterol | 3.54E-84 | 3.29E-86 | NA | 0.17 | 0.15 | 0.18 | 0.0084 | Quantitative Measure | Blood Biochemistry | + |
| Q0504 | Apolipoprotein A | 8.44E-75 | 8.27E-77 | NA | 0.16 | 0.14 | 0.17 | 0.0084 | Quantitative Measure | Blood Biochemistry | + |
| Q0524 | Sex hormone binding globulin | 2.39E-74 | 2.47E-76 | NA | 0.16 | 0.14 | 0.17 | 0.0084 | Quantitative Measure | Blood Biochemistry | + |
| Q0514 | Gamma glutamyltransferase | 1.54E-67 | 1.67E-69 | NA | -0.14 | -0.16 | -0.13 | 0.008 | Quantitative Measure | Blood Biochemistry | - |
| Q0703 | Creatinine (enzymatic) in urine | 2.31E-60 | 2.62E-62 | NA | -0.13 | -0.15 | -0.12 | 0.008 | Quantitative Measure | Urine assay | - |
| Q0503 | Alanine aminotransferase | 8.51E-58 | 1.01E-59 | NA | -0.13 | -0.15 | -0.12 | 0.008 | Quantitative Measure | Blood Biochemistry | - |
| Q0610 | Platelet crit | 2.49E-55 | 3.08E-57 | NA | 0.13 | 0.11 | 0.14 | 0.008 | Quantitative Measure | Blood Count | + |
| Q1091 | Creatinine | 5.31E-45 | 6.84E-47 | NA | -0.23 | -0.26 | -0.2 | 0.016 | Metabolomics | Metabolomics | - |
| Q0525 | Total bilirubin | 1.19E-43 | 1.59E-45 | NA | -0.11 | -0.13 | -0.098 | 0.0081 | Quantitative Measure | Blood Biochemistry | - |
| Q0609 | Platelet count | 6.82E-39 | 9.50E-41 | NA | 0.11 | 0.091 | 0.12 | 0.008 | Quantitative Measure | Blood Count | + |
| Q0506 | Aspartate aminotransferase | 6.09E-38 | 8.80E-40 | NA | -0.11 | -0.12 | -0.091 | 0.0081 | Quantitative Measure | Blood Biochemistry | - |
| Q0707 | Sodium in urine | 1.36E-34 | 2.04E-36 | NA | -0.1 | -0.12 | -0.085 | 0.008 | Quantitative Measure | Urine assay | - |
| Q1123 | Free Cholesterol to Total Lipids in Very Large HDL percentage | 8.21E-31 | 1.27E-32 | NA | -0.18 | -0.21 | -0.15 | 0.015 | Metabolomics | Metabolomics | - |
| Q0507 | Direct bilirubin | 1.86E-30 | 2.98E-32 | NA | -0.1 | -0.12 | -0.085 | 0.0087 | Quantitative Measure | Blood Biochemistry | - |
| Q0009 | Whole body fat mass | 2.36E-29 | 3.89E-31 | NA | 0.092 | 0.077 | 0.11 | 0.0079 | Quantitative Measure | Anthropometry | + |
| Q1152 | Phospholipids in Large HDL | 2.79E-29 | 4.74E-31 | NA | 0.18 | 0.15 | 0.21 | 0.015 | Metabolomics | Metabolomics | + |
| Q1199 | Total Lipids in Large HDL | 6.73E-29 | 1.18E-30 | NA | 0.18 | 0.15 | 0.21 | 0.016 | Metabolomics | Metabolomics | + |
| Q1099 | Free Cholesterol in Large HDL | 1.30E-28 | 2.34E-30 | NA | 0.18 | 0.15 | 0.21 | 0.015 | Metabolomics | Metabolomics | + |
| Q1078 | Concentration of Large HDL Particles | 2.68E-28 | 4.98E-30 | NA | 0.18 | 0.15 | 0.21 | 0.016 | Metabolomics | Metabolomics | + |
| Q1015 | Cholesterol in Large HDL | 4.70E-28 | 8.97E-30 | NA | 0.17 | 0.14 | 0.2 | 0.015 | Metabolomics | Metabolomics | + |
| Q1010 | Average Diameter for HDL Particles | 5.26E-28 | 1.03E-29 | NA | 0.18 | 0.15 | 0.21 | 0.016 | Metabolomics | Metabolomics | + |
| Q0522 | Phosphate | 2.73E-27 | 5.49E-29 | NA | 0.094 | 0.077 | 0.11 | 0.0084 | Quantitative Measure | Blood Biochemistry | + |
| Q1045 | Cholesteryl Esters in Large HDL | 4.21E-27 | 8.68E-29 | NA | 0.17 | 0.14 | 0.2 | 0.015 | Metabolomics | Metabolomics | + |
| Q1055 | Cholesteryl Esters in Very Large HDL | 3.79E-25 | 8.17E-27 | NA | 0.17 | 0.13 | 0.2 | 0.015 | Metabolomics | Metabolomics | + |
| Q1162 | Phospholipids in Very Large HDL | 3.79E-25 | 8.20E-27 | NA | 0.17 | 0.14 | 0.2 | 0.015 | Metabolomics | Metabolomics | + |
| Q1096 | Free Cholesterol in HDL | 2.02E-24 | 4.48E-26 | NA | 0.16 | 0.13 | 0.2 | 0.016 | Metabolomics | Metabolomics | + |
| Q1210 | Total Lipids in Very Large HDL | 2.37E-24 | 5.37E-26 | NA | 0.16 | 0.13 | 0.19 | 0.015 | Metabolomics | Metabolomics | + |
| Q1117 | Free Cholesterol to Total Lipids in Medium HDL percentage | 1.12E-23 | 2.60E-25 | NA | 0.16 | 0.13 | 0.19 | 0.015 | Metabolomics | Metabolomics | + |
| Q1176 | Phospholipids to Total Lipids in Very Large HDL percentage | 4.10E-23 | 9.73E-25 | NA | 0.16 | 0.13 | 0.19 | 0.015 | Metabolomics | Metabolomics | + |
| Q1130 | HDL Cholesterol | 7.26E-23 | 1.76E-24 | NA | 0.16 | 0.13 | 0.19 | 0.016 | Metabolomics | Metabolomics | + |
| Q1196 | Total Lipids in HDL | 1.89E-22 | 4.69E-24 | NA | 0.16 | 0.13 | 0.19 | 0.016 | Metabolomics | Metabolomics | + |
| Q1088 | Concentration of Very Large HDL Particles | 2.19E-22 | 5.53E-24 | NA | 0.16 | 0.13 | 0.19 | 0.016 | Metabolomics | Metabolomics | + |
| Q1024 | Cholesterol in Very Large HDL | 5.70E-22 | 1.47E-23 | NA | 0.15 | 0.12 | 0.18 | 0.015 | Metabolomics | Metabolomics | + |
| Q1042 | Cholesteryl Esters in HDL | 6.49E-22 | 1.73E-23 | NA | 0.16 | 0.13 | 0.19 | 0.016 | Metabolomics | Metabolomics | + |
| Q0614 | Monocyte count | 6.49E-22 | 1.74E-23 | NA | -0.08 | -0.095 | -0.064 | 0.008 | Quantitative Measure | Blood Count | - |
| Q1149 | Phospholipids in HDL | 9.04E-22 | 2.47E-23 | NA | 0.16 | 0.12 | 0.19 | 0.016 | Metabolomics | Metabolomics | + |
| P2072 | Chronic dry cough | 2.87E-20 | 1.48E-23 | 1.7727 | NA | 1.5845 | 1.983 | 1.1 | Symptoms | Symptoms | + |
| Q0528 | Triglycerides | 3.07E-20 | 8.55E-22 | NA | -0.077 | -0.093 | -0.061 | 0.0081 | Quantitative Measure | Blood Biochemistry | - |
| Q1102 | Free Cholesterol in Medium HDL | 3.48E-20 | 9.86E-22 | NA | 0.15 | 0.12 | 0.18 | 0.015 | Metabolomics | Metabolomics | + |
| P174 | Breast cancer | 2.46E-19 | 2.54E-22 | 1.4216 | NA | 1.3242 | 1.5261 | 1 | Neoplasms | Neoplasms | + |
| Q1007 | Apolipoprotein A1 | 3.98E-18 | 1.17E-19 | NA | 0.14 | 0.11 | 0.17 | 0.016 | Metabolomics | Metabolomics | + |
| Q1018 | Cholesterol in Medium HDL | 3.98E-18 | 1.15E-19 | NA | 0.14 | 0.11 | 0.17 | 0.015 | Metabolomics | Metabolomics | + |
| Q1081 | Concentration of Medium HDL Particles | 1.48E-17 | 4.51E-19 | NA | 0.14 | 0.11 | 0.17 | 0.016 | Metabolomics | Metabolomics | + |
| Q0510 | Cholesterol | 1.48E-17 | 4.49E-19 | NA | 0.072 | 0.056 | 0.088 | 0.008 | Quantitative Measure | Blood Biochemistry | + |
| Q0521 | Oestradiol | 1.63E-17 | 5.04E-19 | NA | 0.18 | 0.14 | 0.21 | 0.02 | Quantitative Measure | Blood Biochemistry | + |
| Q1048 | Cholesteryl Esters in Medium HDL | 1.96E-17 | 6.16E-19 | NA | 0.14 | 0.11 | 0.17 | 0.015 | Metabolomics | Metabolomics | + |
| Q1203 | Total Lipids in Medium HDL | 1.49E-16 | 4.78E-18 | NA | 0.14 | 0.1 | 0.17 | 0.016 | Metabolomics | Metabolomics | + |
| Q0207 | Total BMC (bone mineral content) | 1.62E-16 | 5.25E-18 | NA | -0.23 | -0.28 | -0.18 | 0.026 | Quantitative Measure | Imaging | - |
| Q1114 | Free Cholesterol to Total Lipids in Large HDL percentage | 2.85E-16 | 9.55E-18 | NA | 0.13 | 0.1 | 0.16 | 0.015 | Metabolomics | Metabolomics | + |
| Q1128 | Glycine | 2.85E-16 | 9.43E-18 | NA | 0.13 | 0.1 | 0.16 | 0.015 | Metabolomics | Metabolomics | + |
| Q1155 | Phospholipids in Medium HDL | 1.15E-15 | 3.92E-17 | NA | 0.13 | 0.099 | 0.16 | 0.015 | Metabolomics | Metabolomics | + |
| Q1038 | Cholesterol to Total Lipids in Very Large HDL percentage | 4.95E-15 | 1.71E-16 | NA | -0.13 | -0.16 | -0.097 | 0.015 | Metabolomics | Metabolomics | - |
| Q1109 | Free Cholesterol in Very Large HDL | 5.22E-15 | 1.83E-16 | NA | 0.13 | 0.097 | 0.16 | 0.015 | Metabolomics | Metabolomics | + |
| Q0508 | Urea | 7.90E-15 | 2.81E-16 | NA | -0.066 | -0.082 | -0.05 | 0.008 | Quantitative Measure | Blood Biochemistry | - |
| P5107 | Cough on most days | 1.23E-14 | 1.90E-17 | 1.4502 | NA | 1.331 | 1.5799 | 1 | Respiratory | Respiratory | + |
| Q1232 | Triglycerides to Phosphoglycerides ratio | 1.31E-14 | 4.74E-16 | NA | -0.12 | -0.15 | -0.095 | 0.015 | Metabolomics | Metabolomics | - |
| P550.1 | Inguinal hernia | 2.86E-14 | 5.90E-17 | 0.75189 | NA | 0.7033 | 0.80382 | 1 | Digestive | Digestive | - |
| Q0612 | Platelet distribution width | 4.59E-14 | 1.68E-15 | NA | -0.064 | -0.079 | -0.048 | 0.008 | Quantitative Measure | Blood Count | - |
| P411 | Ischemic Heart Disease | 5.59E-14 | 1.73E-16 | 0.82211 | NA | 0.78468 | 0.86132 | 1 | Circulatory system | Circulatory system | - |
| P411.4 | Coronary atherosclerosis | 5.59E-14 | 1.66E-16 | 0.78924 | NA | 0.74605 | 0.83491 | 1 | Circulatory system | Circulatory system | - |
| Q1012 | Average Diameter for VLDL Particles | 1.55E-13 | 5.75E-15 | NA | -0.12 | -0.15 | -0.091 | 0.016 | Metabolomics | Metabolomics | - |
| Q1146 | Phosphatidylcholines | 3.56E-13 | 1.34E-14 | NA | 0.12 | 0.088 | 0.15 | 0.015 | Metabolomics | Metabolomics | + |
| Q1135 | Leucine | 3.77E-13 | 1.44E-14 | NA | -0.12 | -0.15 | -0.088 | 0.015 | Metabolomics | Metabolomics | - |
| Q0200 | Heel bone mineral density (BMD) (lowest) | 5.25E-13 | 2.03E-14 | NA | -0.095 | -0.12 | -0.071 | 0.012 | Quantitative Measure | Imaging | - |
| Q1235 | Triglycerides to Total Lipids in Large HDL percentage | 6.58E-13 | 2.58E-14 | NA | -0.12 | -0.15 | -0.087 | 0.015 | Metabolomics | Metabolomics | - |
| Q1186 | Sphingomyelins | 1.03E-12 | 4.10E-14 | NA | 0.12 | 0.086 | 0.15 | 0.015 | Metabolomics | Metabolomics | + |
| Q1029 | Cholesterol to Total Lipids in Large HDL percentage | 1.11E-12 | 4.45E-14 | NA | 0.12 | 0.086 | 0.15 | 0.015 | Metabolomics | Metabolomics | + |
| Q1075 | Concentration of HDL Particles | 1.16E-12 | 4.71E-14 | NA | 0.12 | 0.087 | 0.15 | 0.016 | Metabolomics | Metabolomics | + |
| Q1189 | Total Cholines | 1.48E-12 | 6.11E-14 | NA | 0.12 | 0.085 | 0.15 | 0.015 | Metabolomics | Metabolomics | + |
| P274 | Gout and other crystal arthropathies | 4.27E-12 | 1.58E-14 | 0.6639 | NA | 0.59798 | 0.73702 | 1.1 | Endocrine/Metabolic | Endocrine/Metabolic | - |
| P274.1 | Gout | 4.27E-12 | 1.76E-14 | 0.64873 | NA | 0.58075 | 0.72458 | 1.1 | Endocrine/Metabolic | Endocrine/Metabolic | - |
| Q1191 | Total Concentration of Lipoprotein Particles | 6.34E-12 | 2.65E-13 | NA | 0.11 | 0.083 | 0.14 | 0.016 | Metabolomics | Metabolomics | + |
| Q0705 | Potassium in urine | 1.07E-11 | 4.51E-13 | NA | -0.058 | -0.073 | -0.042 | 0.008 | Quantitative Measure | Urine assay | - |
| Q1092 | Degree of Unsaturation | 1.31E-11 | 5.62E-13 | NA | 0.11 | 0.081 | 0.14 | 0.015 | Metabolomics | Metabolomics | + |
| Q1147 | Phosphoglycerides | 3.83E-11 | 1.66E-12 | NA | 0.11 | 0.078 | 0.14 | 0.015 | Metabolomics | Metabolomics | + |
| Q1071 | Cholesteryl Esters to Total Lipids in Very Small VLDL percentage | 7.64E-11 | 3.35E-12 | NA | 0.11 | 0.078 | 0.14 | 0.016 | Metabolomics | Metabolomics | + |
| P411.8 | Other chronic ischemic heart disease, unspecified | 1.31E-10 | 6.09E-13 | 0.79686 | NA | 0.74908 | 0.84767 | 1 | Circulatory system | Circulatory system | - |
| Q1241 | Triglycerides to Total Lipids in Small HDL percentage | 8.73E-10 | 3.87E-11 | NA | -0.1 | -0.13 | -0.071 | 0.015 | Metabolomics | Metabolomics | - |
| Q1173 | Phospholipids to Total Lipids in Small HDL percentage | 1.10E-09 | 4.92E-11 | NA | 0.1 | 0.072 | 0.13 | 0.016 | Metabolomics | Metabolomics | + |
| Q0620 | Reticulocyte count | 1.30E-09 | 5.92E-11 | NA | -0.053 | -0.068 | -0.037 | 0.008 | Quantitative Measure | Blood Count | - |
| Q1190 | Total Concentration of Branched-Chain Amino Acids (Leucine + Isoleucine + Valine) | 1.31E-09 | 6.01E-11 | NA | -0.1 | -0.13 | -0.07 | 0.015 | Metabolomics | Metabolomics | - |
| Q1040 | Cholesterol to Total Lipids in Very Small VLDL percentage | 1.36E-09 | 6.33E-11 | NA | 0.1 | 0.071 | 0.13 | 0.016 | Metabolomics | Metabolomics | + |
| Q1093 | Docosahexaenoic Acid | 1.40E-09 | 6.55E-11 | NA | 0.1 | 0.07 | 0.13 | 0.015 | Metabolomics | Metabolomics | + |
| Q1062 | Cholesteryl Esters to Total Lipids in Large VLDL percentage | 2.07E-09 | 9.81E-11 | NA | 0.1 | 0.07 | 0.13 | 0.015 | Metabolomics | Metabolomics | + |
| Q1213 | Total Phospholipids in Lipoprotein Particles | 3.63E-09 | 1.74E-10 | NA | 0.1 | 0.069 | 0.13 | 0.016 | Metabolomics | Metabolomics | + |
| Q1246 | Triglycerides to Total Lipids in Very Small VLDL percentage | 5.80E-09 | 2.81E-10 | NA | -0.098 | -0.13 | -0.068 | 0.016 | Metabolomics | Metabolomics | - |
| Q1034 | Cholesterol to Total Lipids in Medium VLDL percentage | 6.16E-09 | 3.02E-10 | NA | 0.097 | 0.067 | 0.13 | 0.015 | Metabolomics | Metabolomics | + |
| Q1065 | Cholesteryl Esters to Total Lipids in Medium VLDL percentage | 6.54E-09 | 3.24E-10 | NA | 0.097 | 0.067 | 0.13 | 0.015 | Metabolomics | Metabolomics | + |
| Q1170 | Phospholipids to Total Lipids in Medium HDL percentage | 6.72E-09 | 3.36E-10 | NA | -0.097 | -0.13 | -0.066 | 0.015 | Metabolomics | Metabolomics | - |
| Q1120 | Free Cholesterol to Total Lipids in Small HDL percentage | 8.23E-09 | 4.16E-10 | NA | 0.097 | 0.067 | 0.13 | 0.016 | Metabolomics | Metabolomics | + |
| P735.3 | Hallux valgus (Bunion) | 8.59E-09 | 4.43E-11 | 1.4179 | NA | 1.278 | 1.573 | 1.1 | Musculoskeletal | Musculoskeletal | + |
| P411.2 | Myocardial infarction | 9.17E-09 | 5.20E-11 | 0.79441 | NA | 0.74164 | 0.8509 | 1 | Safety | Safety | - |
| Q1060 | Cholesteryl Esters to Total Lipids in Large HDL percentage | 9.64E-09 | 4.92E-10 | NA | 0.096 | 0.066 | 0.13 | 0.015 | Metabolomics | Metabolomics | + |
| Q1240 | Triglycerides to Total Lipids in Medium VLDL percentage | 1.27E-08 | 6.54E-10 | NA | -0.095 | -0.13 | -0.065 | 0.015 | Metabolomics | Metabolomics | - |
| Q1032 | Cholesterol to Total Lipids in Medium HDL percentage | 1.32E-08 | 6.88E-10 | NA | 0.095 | 0.065 | 0.12 | 0.015 | Metabolomics | Metabolomics | + |
| P2013 | Myocardial infarction | 1.46E-08 | 9.03E-11 | 0.79842 | NA | 0.74587 | 0.85464 | 1 | Safety | Safety | - |
| Q0607 | Mean corpuscular haemoglobin concentration | 1.69E-08 | 8.90E-10 | NA | -0.049 | -0.065 | -0.033 | 0.008 | Quantitative Measure | Blood Count | - |
| Q1230 | Triglycerides in Very Large VLDL | 2.16E-08 | 1.15E-09 | NA | -0.095 | -0.13 | -0.064 | 0.016 | Metabolomics | Metabolomics | - |
| Q0100 | Diastolic blood pressure | 3.67E-08 | 1.97E-09 | NA | -0.048 | -0.064 | -0.033 | 0.0081 | Quantitative Measure | Body Measurements | - |
| Q1215 | Triglycerides in Chylomicrons and Extremely Large VLDL | 4.03E-08 | 2.18E-09 | NA | -0.096 | -0.13 | -0.065 | 0.016 | Metabolomics | Metabolomics | - |
| P2210 | Migraine (primary care) | 4.43E-08 | 2.97E-10 | 1.1854 | NA | 1.1243 | 1.2498 | 1 | Neurological | Neurological | + |
| Q1094 | Docosahexaenoic Acid to Total Fatty Acids percentage | 4.65E-08 | 2.54E-09 | NA | 0.092 | 0.061 | 0.12 | 0.015 | Metabolomics | Metabolomics | + |
| Q1211 | Total Lipids in Very Large VLDL | 4.82E-08 | 2.66E-09 | NA | -0.092 | -0.12 | -0.062 | 0.015 | Metabolomics | Metabolomics | - |
| Q1089 | Concentration of Very Large VLDL Particles | 6.37E-08 | 3.55E-09 | NA | -0.092 | -0.12 | -0.062 | 0.016 | Metabolomics | Metabolomics | - |
| Q1057 | Cholesteryl Esters in Very Small VLDL | 9.18E-08 | 5.16E-09 | NA | 0.091 | 0.061 | 0.12 | 0.016 | Metabolomics | Metabolomics | + |
| Q0004 | Body mass index | 9.27E-08 | 5.26E-09 | NA | -0.046 | -0.062 | -0.031 | 0.0079 | Quantitative Measure | Anthropometry | - |
| Q1039 | Cholesterol to Total Lipids in Very Large VLDL percentage | 1.09E-07 | 6.28E-09 | NA | 0.091 | 0.06 | 0.12 | 0.016 | Metabolomics | Metabolomics | + |
| Q0206 | Total BMD (bone mineral density) | 1.09E-07 | 6.24E-09 | NA | -0.15 | -0.2 | -0.1 | 0.026 | Quantitative Measure | Imaging | - |
| Q1070 | Cholesteryl Esters to Total Lipids in Very Large VLDL percentage | 1.13E-07 | 6.59E-09 | NA | 0.092 | 0.061 | 0.12 | 0.016 | Metabolomics | Metabolomics | + |
| Q1119 | Free Cholesterol to Total Lipids in Medium VLDL percentage | 1.13E-07 | 6.62E-09 | NA | 0.089 | 0.059 | 0.12 | 0.015 | Metabolomics | Metabolomics | + |
| Q1238 | Triglycerides to Total Lipids in Medium HDL percentage | 1.34E-07 | 7.93E-09 | NA | -0.089 | -0.12 | -0.059 | 0.015 | Metabolomics | Metabolomics | - |
| Q1245 | Triglycerides to Total Lipids in Very Large VLDL percentage | 1.47E-07 | 8.78E-09 | NA | -0.09 | -0.12 | -0.059 | 0.016 | Metabolomics | Metabolomics | - |
| Q1115 | Free Cholesterol to Total Lipids in Large LDL percentage | 1.51E-07 | 9.20E-09 | NA | 0.088 | 0.058 | 0.12 | 0.015 | Metabolomics | Metabolomics | + |
| Q0210 | Pulse wave Arterial stiffness index | 1.51E-07 | 9.13E-09 | NA | -0.078 | -0.1 | -0.052 | 0.014 | Quantitative Measure | Body Measurements | - |
| Q1180 | Polyunsaturated Fatty Acids to Monounsaturated Fatty Acids ratio | 1.58E-07 | 9.72E-09 | NA | 0.088 | 0.058 | 0.12 | 0.015 | Metabolomics | Metabolomics | + |
| Q1195 | Total Lipids in Chylomicrons and Extremely Large VLDL | 1.58E-07 | 9.79E-09 | NA | -0.089 | -0.12 | -0.059 | 0.016 | Metabolomics | Metabolomics | - |
| Q1139 | Monounsaturated Fatty Acids to Total Fatty Acids percentage | 1.91E-07 | 1.19E-08 | NA | -0.088 | -0.12 | -0.058 | 0.015 | Metabolomics | Metabolomics | - |
| P596.1 | Bladder neck obstruction | 1.94E-07 | 1.40E-09 | 0.55861 | NA | 0.46257 | 0.6744 | 1.1 | Genitourinary | Genitourinary | - |
| P735 | Acquired foot deformities | 2.12E-07 | 1.64E-09 | 1.3011 | NA | 1.1944 | 1.4173 | 1 | Musculoskeletal | Musculoskeletal | + |
| Q1249 | Valine | 2.24E-07 | 1.41E-08 | NA | -0.087 | -0.12 | -0.057 | 0.015 | Metabolomics | Metabolomics | - |
| Q1074 | Concentration of Chylomicrons and Extremely Large VLDL Particles | 2.47E-07 | 1.57E-08 | NA | -0.088 | -0.12 | -0.058 | 0.016 | Metabolomics | Metabolomics | - |
| P2073.1 | Irritable Bowel Syndrome (whole pop controls) | 2.75E-07 | 2.41E-09 | 1.1459 | NA | 1.0958 | 1.1983 | 1 | Digestive | Digestive | + |
| P743.1 | Osteoporosis | 2.75E-07 | 2.37E-09 | 1.2853 | NA | 1.1836 | 1.3957 | 1 | Musculoskeletal | Musculoskeletal | + |
| P743.11 | Osteoporosis NOS | 2.81E-07 | 2.61E-09 | 1.2845 | NA | 1.1829 | 1.3948 | 1 | Musculoskeletal | Musculoskeletal | + |
| Q1112 | Free Cholesterol to Total Lipids in Chylomicrons and Extremely Large VLDL percentage | 3.06E-07 | 1.96E-08 | NA | 0.098 | 0.064 | 0.13 | 0.017 | Metabolomics | Metabolomics | + |
| Q0203 | Visceral adipose tissue volume (VAT) | 3.43E-07 | 2.21E-08 | NA | -0.19 | -0.25 | -0.12 | 0.033 | Quantitative Measure | Imaging | - |
| Q1043 | Cholesteryl Esters in IDL | 3.46E-07 | 2.25E-08 | NA | 0.087 | 0.057 | 0.12 | 0.016 | Metabolomics | Metabolomics | + |
| Q1163 | Phospholipids in Very Large VLDL | 5.10E-07 | 3.34E-08 | NA | -0.089 | -0.12 | -0.058 | 0.016 | Metabolomics | Metabolomics | - |
| Q1014 | Cholesterol in IDL | 5.47E-07 | 3.61E-08 | NA | 0.086 | 0.055 | 0.12 | 0.016 | Metabolomics | Metabolomics | + |
| Q1101 | Free Cholesterol in Large VLDL | 6.70E-07 | 4.46E-08 | NA | -0.085 | -0.12 | -0.055 | 0.016 | Metabolomics | Metabolomics | - |
| Q1219 | Triglycerides in Large HDL | 6.89E-07 | 4.62E-08 | NA | 0.084 | 0.054 | 0.11 | 0.015 | Metabolomics | Metabolomics | + |
| Q1124 | Free Cholesterol to Total Lipids in Very Large VLDL percentage | 7.21E-07 | 4.87E-08 | NA | 0.091 | 0.059 | 0.12 | 0.017 | Metabolomics | Metabolomics | + |
| Q1150 | Phospholipids in IDL | 8.64E-07 | 5.88E-08 | NA | 0.085 | 0.054 | 0.12 | 0.016 | Metabolomics | Metabolomics | + |
| Q1221 | Triglycerides in Large VLDL | 1.10E-06 | 7.55E-08 | NA | -0.083 | -0.11 | -0.053 | 0.015 | Metabolomics | Metabolomics | - |
| Q1080 | Concentration of Large VLDL Particles | 1.11E-06 | 7.71E-08 | NA | -0.084 | -0.11 | -0.053 | 0.016 | Metabolomics | Metabolomics | - |
| Q1167 | Phospholipids to Total Lipids in Large HDL percentage | 1.11E-06 | 7.69E-08 | NA | -0.083 | -0.11 | -0.052 | 0.015 | Metabolomics | Metabolomics | - |
| Q1197 | Total Lipids in IDL | 1.20E-06 | 8.40E-08 | NA | 0.084 | 0.053 | 0.11 | 0.016 | Metabolomics | Metabolomics | + |
| Q1169 | Phospholipids to Total Lipids in Large VLDL percentage | 1.35E-06 | 9.56E-08 | NA | -0.083 | -0.11 | -0.053 | 0.016 | Metabolomics | Metabolomics | - |
| Q1192 | Total Esterified Cholesterol | 1.39E-06 | 9.88E-08 | NA | 0.083 | 0.053 | 0.11 | 0.016 | Metabolomics | Metabolomics | + |
| Q0616 | Eosinophil count | 1.42E-06 | 1.02E-07 | NA | -0.042 | -0.057 | -0.027 | 0.0079 | Quantitative Measure | Blood Count | - |
| Q1201 | Total Lipids in Large VLDL | 1.50E-06 | 1.08E-07 | NA | -0.082 | -0.11 | -0.052 | 0.015 | Metabolomics | Metabolomics | - |
| P2057 | Ever smoked | 1.80E-06 | 1.76E-08 | 0.90917 | NA | 0.87955 | 0.93979 | 1 | Behaviour | Behaviour | - |
| Q1175 | Phospholipids to Total Lipids in Small VLDL percentage | 1.97E-06 | 1.43E-07 | NA | 0.082 | 0.052 | 0.11 | 0.016 | Metabolomics | Metabolomics | + |
| Q1122 | Free Cholesterol to Total Lipids in Small VLDL percentage | 2.06E-06 | 1.51E-07 | NA | 0.081 | 0.051 | 0.11 | 0.015 | Metabolomics | Metabolomics | + |
| Q1026 | Cholesterol in Very Small VLDL | 2.29E-06 | 1.69E-07 | NA | 0.082 | 0.051 | 0.11 | 0.016 | Metabolomics | Metabolomics | + |
| Q1181 | Polyunsaturated Fatty Acids to Total Fatty Acids percentage | 2.72E-06 | 2.02E-07 | NA | 0.08 | 0.05 | 0.11 | 0.015 | Metabolomics | Metabolomics | + |
| Q1228 | Triglycerides in VLDL | 2.79E-06 | 2.10E-07 | NA | -0.081 | -0.11 | -0.05 | 0.016 | Metabolomics | Metabolomics | - |
| Q0606 | Mean corpuscular haemoglobin | 2.79E-06 | 2.10E-07 | NA | -0.041 | -0.057 | -0.026 | 0.008 | Quantitative Measure | Blood Count | - |
| P5000 | Snoring | 2.84E-06 | 2.93E-08 | 0.91063 | NA | 0.881 | 0.94126 | 1 | Symptoms | Sleep | - |
| Q1132 | Isoleucine | 3.11E-06 | 2.36E-07 | NA | -0.081 | -0.11 | -0.05 | 0.016 | Metabolomics | Metabolomics | - |
| Q1097 | Free Cholesterol in IDL | 3.16E-06 | 2.41E-07 | NA | 0.081 | 0.05 | 0.11 | 0.016 | Metabolomics | Metabolomics | + |
| Q1225 | Triglycerides in Small HDL | 3.41E-06 | 2.62E-07 | NA | -0.079 | -0.11 | -0.049 | 0.015 | Metabolomics | Metabolomics | - |
| Q1178 | Phospholipids to Total Lipids in Very Small VLDL percentage | 3.72E-06 | 2.88E-07 | NA | -0.08 | -0.11 | -0.049 | 0.016 | Metabolomics | Metabolomics | - |
| Q0625 | High light scatter reticulocyte count | 4.04E-06 | 3.15E-07 | NA | -0.041 | -0.057 | -0.025 | 0.008 | Quantitative Measure | Blood Count | - |
| Q1013 | Cholesterol in Chylomicrons and Extremely Large VLDL | 4.65E-06 | 3.67E-07 | NA | -0.082 | -0.11 | -0.05 | 0.016 | Metabolomics | Metabolomics | - |
| Q1154 | Phospholipids in Large VLDL | 4.65E-06 | 3.67E-07 | NA | -0.08 | -0.11 | -0.049 | 0.016 | Metabolomics | Metabolomics | - |
| Q1187 | Total Cholesterol | 5.77E-06 | 4.58E-07 | NA | 0.079 | 0.048 | 0.11 | 0.016 | Metabolomics | Metabolomics | + |
| Q1027 | Cholesterol to Total Lipids in Chylomicrons and Extremely Large VLDL percentage | 6.08E-06 | 4.86E-07 | NA | 0.081 | 0.05 | 0.11 | 0.016 | Metabolomics | Metabolomics | + |
| Q1172 | Phospholipids to Total Lipids in Medium VLDL percentage | 7.27E-06 | 5.85E-07 | NA | 0.077 | 0.047 | 0.11 | 0.015 | Metabolomics | Metabolomics | + |
| P565 | Anal and rectal conditions | 7.48E-06 | 8.10E-08 | 0.83685 | NA | 0.78412 | 0.8931 | 1 | Digestive | Digestive | - |
| Q1025 | Cholesterol in Very Large VLDL | 1.13E-05 | 9.18E-07 | NA | -0.076 | -0.11 | -0.046 | 0.016 | Metabolomics | Metabolomics | - |
| P317 | Alcohol-related disorders | 1.28E-05 | 1.52E-07 | 0.83627 | NA | 0.78226 | 0.89397 | 1 | Psychiatric disorders | Psychiatric disorders | - |
| P599.4 | Urinary incontinence | 1.28E-05 | 1.48E-07 | 1.2556 | NA | 1.1534 | 1.3668 | 1 | Genitourinary | Genitourinary | + |
| P2073 | Irritable Bowel Syndrome | 1.30E-05 | 1.61E-07 | 1.1466 | NA | 1.0894 | 1.2068 | 1 | Digestive | Digestive | + |
| Q1148 | Phospholipids in Chylomicrons and Extremely Large VLDL | 1.55E-05 | 1.26E-06 | NA | -0.083 | -0.12 | -0.05 | 0.017 | Metabolomics | Metabolomics | - |
| Q0011 | birth weight | 1.67E-05 | 1.37E-06 | NA | -0.049 | -0.07 | -0.029 | 0.01 | Quantitative Measure | Anthropometry | - |
| P2021 | Acute urinary retention | 1.89E-05 | 2.46E-07 | 0.80856 | NA | 0.74584 | 0.8765 | 1 | Safety | Safety | - |
| P599.2 | Retention of urine | 1.89E-05 | 2.54E-07 | 0.81169 | NA | 0.74976 | 0.87868 | 1 | Safety | Safety | - |
| Q0110 | Systolic blood pressure | 1.99E-05 | 1.64E-06 | NA | -0.039 | -0.054 | -0.023 | 0.0081 | Quantitative Measure | Body Measurements | - |
| Q1118 | Free Cholesterol to Total Lipids in Medium LDL percentage | 2.14E-05 | 1.78E-06 | NA | 0.073 | 0.043 | 0.1 | 0.015 | Metabolomics | Metabolomics | + |
| P244.4 | Hypothyroidism NOS | 2.34E-05 | 3.26E-07 | 1.1833 | NA | 1.1093 | 1.2623 | 1 | Endocrine/Metabolic | Endocrine/Metabolic | + |
| Q1234 | Triglycerides to Total Lipids in IDL percentage | 2.67E-05 | 2.23E-06 | NA | -0.074 | -0.1 | -0.043 | 0.016 | Metabolomics | Metabolomics | - |
| P244 | Hypothyroidism | 3.43E-05 | 4.96E-07 | 1.1758 | NA | 1.1039 | 1.2524 | 1 | Endocrine/Metabolic | Endocrine/Metabolic | + |
| Q1009 | Apolipoprotein B to Apolipoprotein A1 ratio | 3.54E-05 | 2.98E-06 | NA | -0.073 | -0.1 | -0.042 | 0.016 | Metabolomics | Metabolomics | - |
| P743 | Osteoporosis, osteopenia and pathological fracture | 3.66E-05 | 5.47E-07 | 1.2175 | NA | 1.1272 | 1.3149 | 1 | Musculoskeletal | Musculoskeletal | + |
| P426 | Cardiac conduction disorders | 3.74E-05 | 5.78E-07 | 0.82357 | NA | 0.76319 | 0.88869 | 1 | Circulatory system | Circulatory system | - |
| Q1011 | Average Diameter for LDL Particles | 3.98E-05 | 3.37E-06 | NA | 0.073 | 0.042 | 0.1 | 0.016 | Metabolomics | Metabolomics | + |
| Q1179 | Polyunsaturated Fatty Acids | 5.66E-05 | 4.82E-06 | NA | 0.07 | 0.04 | 0.1 | 0.015 | Metabolomics | Metabolomics | + |
| Q1214 | Total Triglycerides | 5.74E-05 | 4.91E-06 | NA | -0.071 | -0.1 | -0.041 | 0.016 | Metabolomics | Metabolomics | - |
| Q1059 | Cholesteryl Esters to Total Lipids in IDL percentage | 6.40E-05 | 5.51E-06 | NA | 0.071 | 0.04 | 0.1 | 0.016 | Metabolomics | Metabolomics | + |
| Q1017 | Cholesterol in Large VLDL | 6.58E-05 | 5.70E-06 | NA | -0.07 | -0.1 | -0.04 | 0.015 | Metabolomics | Metabolomics | - |
| Q1141 | Omega-3 Fatty Acids to Total Fatty Acids percentage | 8.44E-05 | 7.36E-06 | NA | 0.069 | 0.039 | 0.099 | 0.015 | Metabolomics | Metabolomics | + |
| P565.1 | Anal and rectal polyp | 8.76E-05 | 1.40E-06 | 0.80877 | NA | 0.74193 | 0.88157 | 1 | Digestive | Digestive | - |
| Q0509 | Calcium | 1.13E-04 | 9.91E-06 | NA | 0.037 | 0.021 | 0.054 | 0.0084 | Quantitative Measure | Blood Biochemistry | + |
| Q0519 | LDL direct | 1.19E-04 | 1.05E-05 | NA | 0.036 | 0.02 | 0.051 | 0.0081 | Quantitative Measure | Blood Biochemistry | + |
| Q0613 | Lymphocyte count | 1.22E-04 | 1.08E-05 | NA | 0.035 | 0.019 | 0.051 | 0.008 | Quantitative Measure | Blood Count | + |
| Q1041 | Cholesteryl Esters in Chylomicrons and Extremely Large VLDL | 1.23E-04 | 1.10E-05 | NA | -0.075 | -0.11 | -0.041 | 0.017 | Metabolomics | Metabolomics | - |
| Q1031 | Cholesterol to Total Lipids in Large VLDL percentage | 1.31E-04 | 1.18E-05 | NA | 0.068 | 0.037 | 0.098 | 0.015 | Metabolomics | Metabolomics | + |
| P2063 | Family history High blood pressure | 1.34E-04 | 2.21E-06 | 1.0773 | NA | 1.0446 | 1.111 | 1 | Circulatory system | Circulatory system | + |
| Q2020 | eGFR | 1.37E-04 | 1.24E-05 | NA | 0.035 | 0.019 | 0.051 | 0.008 | Quantitative Measure | Blood Biochemistry | + |
| Q1058 | Cholesteryl Esters to Total Lipids in Chylomicrons and Extremely Large VLDL percentage | 1.50E-04 | 1.36E-05 | NA | 0.074 | 0.041 | 0.11 | 0.017 | Metabolomics | Metabolomics | + |
| Q1224 | Triglycerides in Medium VLDL | 1.60E-04 | 1.46E-05 | NA | -0.067 | -0.097 | -0.036 | 0.015 | Metabolomics | Metabolomics | - |
| Q1243 | Triglycerides to Total Lipids in Small VLDL percentage | 1.85E-04 | 1.70E-05 | NA | -0.067 | -0.098 | -0.037 | 0.016 | Metabolomics | Metabolomics | - |
| Q1028 | Cholesterol to Total Lipids in IDL percentage | 1.88E-04 | 1.74E-05 | NA | 0.067 | 0.036 | 0.098 | 0.016 | Metabolomics | Metabolomics | + |
| Q1140 | Omega-3 Fatty Acids | 2.04E-04 | 1.89E-05 | NA | 0.066 | 0.036 | 0.096 | 0.015 | Metabolomics | Metabolomics | + |
| Q1063 | Cholesteryl Esters to Total Lipids in Medium HDL percentage | 2.14E-04 | 2.00E-05 | NA | 0.066 | 0.035 | 0.096 | 0.015 | Metabolomics | Metabolomics | + |
| Q1194 | Total Free Cholesterol | 2.43E-04 | 2.28E-05 | NA | 0.066 | 0.036 | 0.097 | 0.016 | Metabolomics | Metabolomics | + |
| Q1242 | Triglycerides to Total Lipids in Small LDL percentage | 2.47E-04 | 2.33E-05 | NA | -0.069 | -0.1 | -0.037 | 0.016 | Metabolomics | Metabolomics | - |
| Q0123 | FEV1/FVC ratio | 3.16E-04 | 3.00E-05 | NA | 0.037 | 0.02 | 0.055 | 0.009 | Quantitative Measure | Spirometry | + |
| P427 | Cardiac dysrhythmias | 3.18E-04 | 5.41E-06 | 0.88735 | NA | 0.8428 | 0.93424 | 1 | Circulatory system | Circulatory system | - |
| Q1095 | Free Cholesterol in Chylomicrons and Extremely Large VLDL | 3.32E-04 | 3.17E-05 | NA | -0.072 | -0.11 | -0.038 | 0.017 | Metabolomics | Metabolomics | - |
| Q1142 | Omega-6 Fatty Acids | 3.69E-04 | 3.54E-05 | NA | 0.064 | 0.033 | 0.094 | 0.015 | Metabolomics | Metabolomics | + |
| P427.2 | Atrial fibrillation and flutter | 3.72E-04 | 6.53E-06 | 0.84714 | NA | 0.78817 | 0.91047 | 1 | Circulatory system | Circulatory system | - |
| P411.3 | Angina pectoris | 3.74E-04 | 6.75E-06 | 0.86225 | NA | 0.80835 | 0.91971 | 1 | Circulatory system | Circulatory system | - |
| Q0204 | Abdominal subcutaneous adipose tissue volume | 3.87E-04 | 3.73E-05 | NA | 0.14 | 0.072 | 0.2 | 0.033 | Quantitative Measure | Imaging | + |
| Q1227 | Triglycerides in Small VLDL | 4.69E-04 | 4.55E-05 | NA | -0.064 | -0.094 | -0.033 | 0.016 | Metabolomics | Metabolomics | - |
| P512.8 | Cough | 5.35E-04 | 9.93E-06 | 1.3239 | NA | 1.1689 | 1.4992 | 1.1 | Respiratory | Respiratory | + |
| Q1110 | Free Cholesterol in Very Large VLDL | 8.96E-04 | 8.73E-05 | NA | -0.066 | -0.099 | -0.033 | 0.017 | Metabolomics | Metabolomics | - |
| P596 | Other disorders of bladder | 1.06E-03 | 2.02E-05 | 0.83914 | NA | 0.7741 | 0.90958 | 1 | Genitourinary | Genitourinary | - |
| Q1030 | Cholesterol to Total Lipids in Large LDL percentage | 1.10E-03 | 1.08E-04 | NA | 0.06 | 0.029 | 0.09 | 0.015 | Metabolomics | Metabolomics | + |
| Q1209 | Total Lipids in VLDL | 1.18E-03 | 1.16E-04 | NA | -0.06 | -0.091 | -0.03 | 0.016 | Metabolomics | Metabolomics | - |
| P306 | Other mental disorder | 1.37E-03 | 2.69E-05 | 0.9126 | NA | 0.87446 | 0.9524 | 1 | Psychiatric disorders | Psychiatric disorders | - |
| P819 | Skull and face fracture and other intercranial injury | 1.52E-03 | 3.06E-05 | 0.74263 | NA | 0.64563 | 0.85405 | 1.1 | Injuries & poisonings | Injuries & poisonings | - |
| Q1056 | Cholesteryl Esters in Very Large VLDL | 1.54E-03 | 1.52E-04 | NA | -0.06 | -0.091 | -0.029 | 0.016 | Metabolomics | Metabolomics | - |
| Q1037 | Cholesterol to Total Lipids in Small VLDL percentage | 1.60E-03 | 1.59E-04 | NA | 0.059 | 0.028 | 0.089 | 0.016 | Metabolomics | Metabolomics | + |
| Q2010 | Blood Potassium (primary care) | 1.85E-03 | 1.85E-04 | NA | -0.047 | -0.072 | -0.022 | 0.013 | Quantitative Measure | Blood Biochemistry | - |
| Q1233 | Triglycerides to Total Lipids in Chylomicrons and Extremely Large VLDL percentage | 2.05E-03 | 2.06E-04 | NA | -0.06 | -0.091 | -0.028 | 0.016 | Metabolomics | Metabolomics | - |
| P611 | Abnormal findings on mammogram or breast exam | 2.19E-03 | 4.51E-05 | 1.5634 | NA | 1.261 | 1.9374 | 1.1 | Genitourinary | Genitourinary | + |
| Q1105 | Free Cholesterol in Small HDL | 2.20E-03 | 2.22E-04 | NA | 0.058 | 0.027 | 0.088 | 0.016 | Metabolomics | Metabolomics | + |
| P272.11 | Hypercholesterolemia | 2.47E-03 | 5.23E-05 | 0.91282 | NA | 0.87335 | 0.95406 | 1 | Endocrine/Metabolic | Endocrine/Metabolic | - |
| Q0502 | Alkaline phosphatase | 2.66E-03 | 2.70E-04 | NA | 0.029 | 0.014 | 0.045 | 0.008 | Quantitative Measure | Blood Biochemistry | + |
| P272.1 | Hyperlipidemia | 3.74E-03 | 8.10E-05 | 0.91754 | NA | 0.8791 | 0.95765 | 1 | Endocrine/Metabolic | Endocrine/Metabolic | - |
| Q1100 | Free Cholesterol in Large LDL | 3.82E-03 | 3.90E-04 | NA | 0.054 | 0.024 | 0.085 | 0.015 | Metabolomics | Metabolomics | + |
| Q1226 | Triglycerides in Small LDL | 3.90E-03 | 4.00E-04 | NA | -0.058 | -0.09 | -0.026 | 0.016 | Metabolomics | Metabolomics | - |
| Q0621 | Mean reticulocyte volume | 3.96E-03 | 4.08E-04 | NA | -0.028 | -0.044 | -0.013 | 0.008 | Quantitative Measure | Blood Count | - |
| Q1111 | Free Cholesterol in Very Small VLDL | 4.04E-03 | 4.19E-04 | NA | 0.055 | 0.024 | 0.086 | 0.016 | Metabolomics | Metabolomics | + |
| P272 | Disorders of lipoid metabolism | 4.17E-03 | 9.40E-05 | 0.9184 | NA | 0.87999 | 0.95848 | 1 | Endocrine/Metabolic | Endocrine/Metabolic | - |
| P594 | Urinary calculus | 4.17E-03 | 9.46E-05 | 0.81877 | NA | 0.74054 | 0.90519 | 1.1 | Genitourinary | Genitourinary | - |
| Q1144 | Omega-6 Fatty Acids to Total Fatty Acids percentage | 4.36E-03 | 4.54E-04 | NA | 0.054 | 0.024 | 0.084 | 0.015 | Metabolomics | Metabolomics | + |
| P426.3 | Bundle branch block | 4.52E-03 | 1.05E-04 | 0.81436 | NA | 0.73404 | 0.90338 | 1.1 | Circulatory system | Circulatory system | - |
| Q0209 | Carotid intima media thickness (IMT) (mean) | 4.62E-03 | 4.85E-04 | NA | -0.084 | -0.13 | -0.037 | 0.024 | Quantitative Measure | Imaging | - |
| P5005 | Pack years | 4.62E-03 | 4.86E-04 | NA | -0.048 | -0.075 | -0.021 | 0.014 | Behaviour | Smoking | - |
| P611.3 | Lump or mass in breast | 5.23E-03 | 1.24E-04 | 1.5367 | NA | 1.2336 | 1.9132 | 1.1 | Genitourinary | Genitourinary | + |
| P550.4 | Umbilical hernia | 6.31E-03 | 1.53E-04 | 0.78149 | NA | 0.6878 | 0.88781 | 1.1 | Digestive | Digestive | - |
| P2035 | Diabetes mellitus | 6.41E-03 | 1.61E-04 | 0.88538 | NA | 0.83112 | 0.94316 | 1 | Endocrine/Metabolic | Endocrine/Metabolic | - |
| P2200.39 | Fibromyalgia pain clinical | 6.41E-03 | 1.62E-04 | 1.3282 | NA | 1.1459 | 1.5391 | 1.1 | Neuropathic Pain | Neuropathic Pain | + |
| Q1212 | Total Lipids in Very Small VLDL | 6.53E-03 | 6.90E-04 | NA | 0.053 | 0.022 | 0.084 | 0.016 | Metabolomics | Metabolomics | + |
| Q1136 | Linoleic Acid | 6.60E-03 | 7.01E-04 | NA | 0.052 | 0.022 | 0.082 | 0.015 | Metabolomics | Metabolomics | + |
| P2035.2 | Type 2 diabetes | 7.25E-03 | 1.87E-04 | 0.88324 | NA | 0.82754 | 0.94267 | 1 | Endocrine/Metabolic | Endocrine/Metabolic | - |
| Q1121 | Free Cholesterol to Total Lipids in Small LDL percentage | 7.52E-03 | 8.03E-04 | NA | 0.052 | 0.021 | 0.082 | 0.015 | Metabolomics | Metabolomics | + |
| P426.31 | Right bundle branch block | 7.57E-03 | 2.03E-04 | 0.75541 | NA | 0.65145 | 0.87579 | 1.1 | Circulatory system | Circulatory system | - |
| P426.9 | Cardiac pacemaker/device in situ | 7.57E-03 | 2.02E-04 | 0.78639 | NA | 0.69274 | 0.89258 | 1.1 | Circulatory system | Circulatory system | - |
| P2076 | Chronic obstructive pulmonary disease (spirometry GOLD1+) | 7.90E-03 | 2.17E-04 | 0.91473 | NA | 0.87252 | 0.95896 | 1 | Respiratory | Respiratory | - |
| P710.11 | Acute osteomyelitis | 7.90E-03 | 2.20E-04 | 0.093031 | NA | 0.026483 | 0.32799 | 1.9 | Musculoskeletal | Musculoskeletal | - |

#### **Supplementary Table 16. Functions of mapped genes.**

| **Gene Symbol** | **Gene Name** | **Description** |
| --- | --- | --- |
| *ALX1* | ALX Homeobox 1 | DNA binding transcription factor likely to regulate expression of genes involved in mesenchyme-derived craniofacial structure development^47^. Associated with Frontonasal Dysplasia 3^48,49^. |
| *ATP23* | ATP23 Metallopeptidase And ATP Synthase Assembly Factor Homolog | Encodes a metallopeptidase involved in DNA repair through binding via the DNA-binding subunit of DNA-dependent protein kinase, and also has a role in mitochondrial protein processing and ATP synthase complex assembly^47^. ATP23 is known to be amplified in glioblastomas^47^, and is highly expressed in the reproductive system^49^. |
| *CPEB2* | Cytoplasmic Polyadenylation Element Binding Protein 2 | Important for transition from metaphase to anaphase in the cell cycle^49^. Involved in regulation of cytoplasmic translation^49^. |
| *CTNNA1* | Catenin alpha 1 | Member of the catenin family which is widely expressed across anatomical systems^49^, and is most well-known for its role in cell-cell adhesion by connecting cadherins to the actin cytoskeleton^47^. Additional functions include cell migration, regulation of apoptosis, axon regeneration and integrin-mediated signalling. CTNNA1 variants have been associated with macular dystrophy (autosomal dominant inheritance)^48^, and other eye diseases including butterfly-shaped pigment dystrophy^49^. |
| *CYP27B1* | Cytochrome P450 Family 27 Subfamily B Member 1 | Member of the cytochrome P450 family localised to the mitochondrial membrane^47,49^. Synthesises the active form of vitamin D3 which is involved in calcium homeostasis^47^. Associated with Vitamin D-dependent rickets, type I^48^. |
| *KCNA10* | Potassium Voltage-Gated Channel Subfamily A Member 10 | Intronless gene encoding a member of the voltage-gated potassium channel complex subfamily^47^. Mediates potassium ion transmembrane transport across plasma membranes^49^. |
| *KCNIP4* | Potassium voltage-gated channel interacting protein 4 | Integral component of voltage-gated potassium channel complexes predominantly expressed in the brain^47,49^. Involved in the regulation of potassium ion transmembrane transport and excitability in neurons and cardomycotyes^50^. Also involved in protein localisation to the plasma membrane^49^. |
| *L3MBTL4* | L3MBTL histone methyl-lysine binding protein 4 | Negatively regulates transcription through chromatin organisation and histone binding activity and has high baseline expression in the immune and hematopoietic systems^47,49^. Following evidence of association between L3MBTL4 and hypertension^51^, in vivo analyses showed that L3MBTL4 induces the proliferation and remodelling of vascular smooth muscle cells via the MAPK pathway, resulting in hypertension^52^. |
| *MAPKAP1* | MAPK associated protein 1 | Subunit of mTORC2 which is involved in a range of biological processes via the mTOR signalling pathway^49^. With widespread expression across anatomical systems, MAPKAP1 regulates cell growth, autophagy, and survival in response to stimuli (e.g. hormones, growth factors and stress)^47^. Dysfunction of mTORC2 has been linked to neurodegenerative disease, epilepsy and cancer^53^. |
| *NTSR1* | Neurotensin receptor 1 | High baseline expression in the nervous system and encodes a G-protein coupled receptor which is implicated in many functions modulated by neurotensin, a vasoactive peptide which binds with high affinity to NTSR1^47,49^. Functions include regulation of neuropeptide signalling pathways, neurotransmitter secretion (specifically, gamma-aminobutyric acid and glutamate) and synaptic transmission^49^. |
| *OR4C12* | Olfactory receptor family 4 subfamily C member 12 | G-protein coupled receptor located in the plasma membrane which is involved in smell perception via the transduction of odorant signals upon detection of specific chemical stimuli^47,49^. Predominantly expressed in the olfactory sensory neurons of the nasal epithelium^54^. |
| *OR4C13* | Olfactory receptor family 4 subfamily C member 13 |  |
| *PREP* | Prolyl endopeptidase | Cytosolic prolyl endopeptidase involved in the maturation and degradation of peptide hormones and neuropeptides^47^, including angiotensin II, bradykinin and oxytocin^55,56^. Has high baseline expression in the digestive system^49^, and is also hypothesised to be involved in the release of Ac-SDKP from its precursor thymosin-β4 which has anti-inflammatory and anti- fibrotic effects^57^. This is supported by the relationship between PREP and inflammatory disease, including neurodegeneration, cancer and COPD^58^. |
| *RASSF9* | Ras Association Domain Family Member 9 | Encoding protein is localised to endosomes and has a role in intracellular and endosomal transport, protein targeting and signal transduction^49^. |
| *RBM15* | RNA Binding Motif Protein 15 | Member of the SPEN family of proteins^47^. Involved in a wide range of biological processes, including RNA methylation, transcription regulation, alternative splicing regulation, and differentiation of myeloid cells and megakaryocytes^49^. |
| *SIL1* | SIL1 nucleotide exchange factor | Glycoprotein which is widely expressed across anatomical systems^49^ and involved in endoplasmic reticulum-based protein translocation and folding^47,49^. Homozygous mutations in SIL1 have been associated with Marinesco-Sjogren syndrome^48^, characterised by ataxia, early onset cataracts, myopathy, muscle weakness, hypotonia and intellectual disability^59^. |
| *SLCO4A1* | Solute Carrier Organic Anion Transporter Family Member 4A1 | Highly expressed in the respiratory system^49^ and located in the plasma membrane^47^. Involved in the transport of various molecules, including prostaglandins, thyroid hormones and organic anions^49^. |
| *SRBD1* | S1 RNA Binding Domain 1 | Likely to be a ribosomal component involved in translation^47,49^. |
| *VMA21* | Vacuolar ATPase assembly factor VMA21 | Chaperone for the assembly of lysosomal vascular ATPase (an enzyme which regulates intracellular pH) and has a high baseline expression in the immune and hematopoietic systems^47,49^. Mutations in VMA21 have been linked to the X-linked recessive disease, myopathy with excessive autophagy^60^, and non-alcoholic fatty liver disease^61^. |

### Supplementary Figures

#### **Supplementary Figure 1. Genetic risk score-based PheWAS results (**false discovery rate [**FDR <0.01]) using DeepPheWAS, restricted to binary phenotypes.**

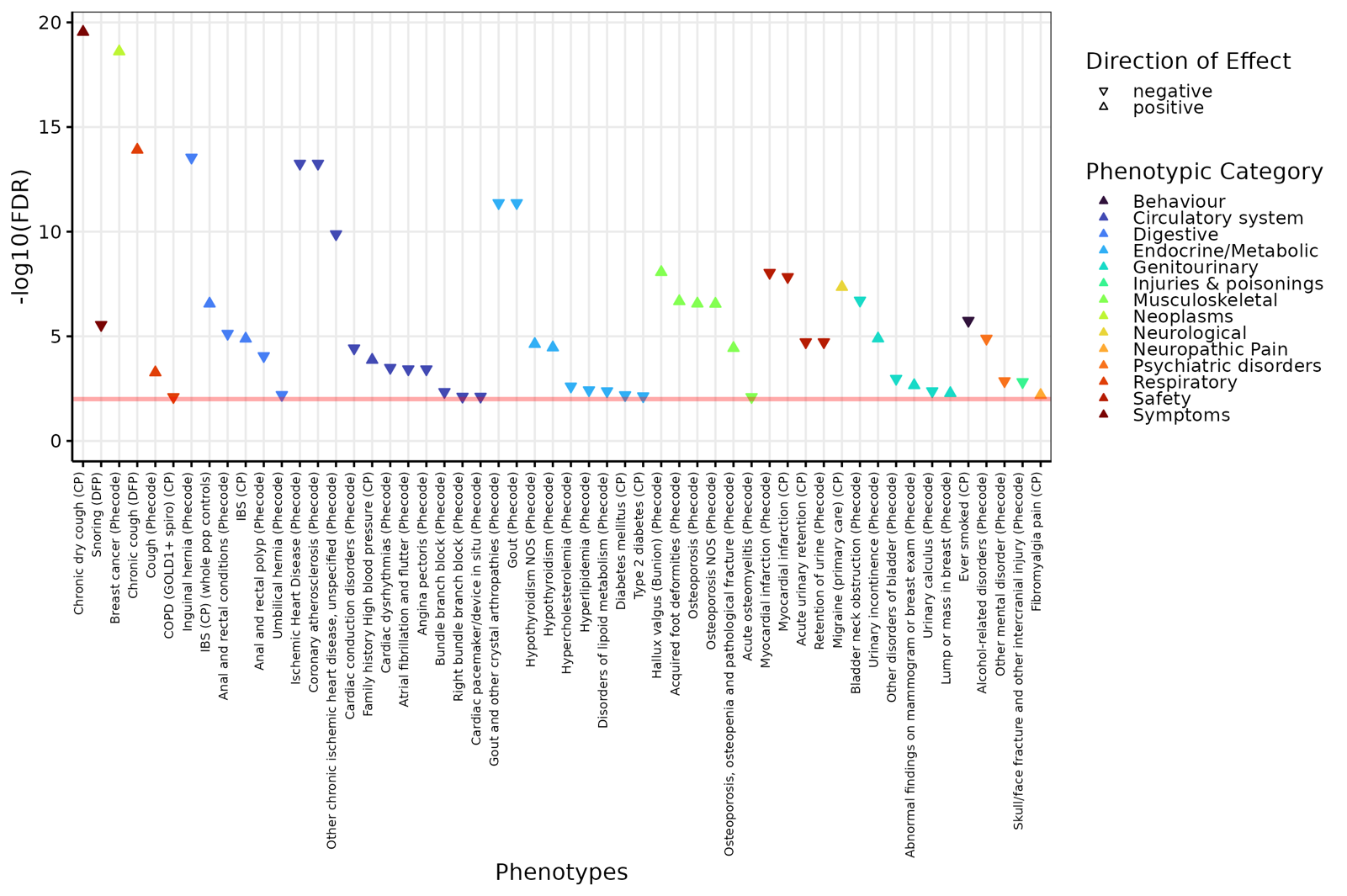

4.	DReichLab. EIG. https://github.com/DReichLab/EIG
